# Association of assisted reproductive technology with long-term offspring cardiometabolic health: a multi-cohort study

**DOI:** 10.1101/2022.04.13.22273455

**Authors:** Ahmed Elhakeem, Amy E Taylor, Hazel M Inskip, Jonathan Huang, Toby Mansell, Carina Rodrigues, Federica Asta, Sophie M Blaauwendraad, Siri E Håberg, Jane Halliday, Margreet W Harskamp-van Ginkel, Jian-Rong He, Vincent WV Jaddoe, Sharon Lewis, Gillian M Maher, Yannis Manios, Fergus P McCarthy, Irwin KM Reiss, Franca Rusconi, Theodosia Salika, Muriel Tafflet, Xiu Qiu, Bjørn O Åsvold, David Burgner, Jerry KY Chan, Luigi Gagliardi, Romy Gaillard, Barbara Heude, Maria C Magnus, George Moschonis, Deirdre Murray, Scott M Nelson, Daniela Porta, Richard Saffery, Henrique Barros, Johan G Eriksson, Tanja GM Vrijkotte, Deborah A Lawlor

**Affiliations:** MRC Integrative Epidemiology Unit at the University of Bristol, Bristol, UK; Population Health Science, Bristol Medical School, University of Bristol, Bristol, UK; NIHR Bristol Biomedical Research Centre, Bristol, UK; MRC Lifecourse Epidemiology Centre, University of Southampton, Southampton, UK; Singapore Institute for Clinical Science, Agency for Science, Technology, and Research, Singapore; Centre for Quantitative Medicine; Duke-NUS Medical School, Singapore; Murdoch Children’s Research Institute, Parkville, VIC, Australia; University of Melbourne, Parkville, VIC, Australia; EPIUnit - Instituto de Saúde Pública, Universidade do Porto, Porto, Portugal; Laboratório para a Investigação Integrativa e Translacional em Saúde Populacional (ITR), Porto, Portugal; Depatment of Epidemiology, Lazio Regional Health Service, Rome, Italy; The Generation R Study Group, Erasmus MC, University Medical Center, Rotterdam, The Netherlands; Department of Paediatrics, Erasmus MC, University Medical Center, Rotterdam, The Netherlands; Centre for Fertility and Health, Norwegian Institute of Public Health, Oslo, Norway; Amsterdam UMC, University of Amsterdam, Department of Public and Occupational Health, Amsterdam Public Health Research Institute, Amsterdam, The Netherlands; Division of Birth Cohort Study, Guangzhou Women and Children’s Medical Center, Guangzhou Medical University, Guangzhou, China; School of Public Health, University College Cork, Cork, Ireland; The Irish Centre for Maternal and Child Health Research (INFANT), University College Cork, Cork, Ireland; Department of Nutrition and Dietetics, School of Health Science and Education, Harokopio University, Athens, Greece; Institute of Agri-Food and Life Sciences, Hellenic Mediterranean University Research Centre, Heraklion, Greece; Department of Obstetrics and Gynaecology, University College Cork, Cork, Ireland; Department of Mother and Child Health, Ospedale Versilia, Viareggio, AUSL Toscana Nord Ovest, Pisa, Italy; Université de Paris, Inserm, INRAE, Centre for Research in Epidemiology and StatisticS (CRESS), Paris, France; K.G. Jebsen Center for Genetic Epidemiology, Department of Public Health and Nursing, Faculty of Medicine and Health Sciences, NTNU, Norwegian University of Science and Technology, Trondheim, Norway; HUNT Research Centre, Department of Public Health and Nursing, Faculty of Medicine and Health Sciences, NTNU, Norwegian University of Science and Technology, Levanger, Norway; Department of Endocrinology, Clinic of Medicine, St. Olavs Hospital, Trondheim University Hospital, Trondheim, Norway; Department of Paediatrics, University of Melbourne, Parkville, VIC, Australia; Department of Paediatrics, Monash University, Clayton, VIC, Australia; Department of Reproductive Medicine, KK Women’s and Children’s Hospital, Singapore; Academic Clinical Program in Obstetrics and Gynaecology, Duke-NUS Medical School, Singapore; Department of Dietetics, Nutrition and Sport, College of Science, Health and Engineering, La Trobe University, Melbourne, Australia; Department of Pediatrics and Child Health, University College Cork, Cork, Ireland; School of Medicine, University of Glasgow, Glasgow, UK; Department of Obstetrics and Gynaecology and Human Potential Translational Research Programme, Yong Loo Lin School of Medicine, National University of Singapore, Singapore; Department of General Practice and Primary Health Care, University of Helsinki and Helsinki University Hospital, Helsinki, Finland; Folkhä lsan Research Center, Helsinki, Finland

**Author notes:** Corresponding author Dr Ahmed Elhakeem, MRC Integrative Epidemiology Unit at the University of Bristol, Bristol, UK; Population Health Science, Bristol Medical School, University of Bristol, Bristol, UK,.

## Abstract

**Objectives:** To examine association of conception by assisted reproductive technology (ART) with offspring cardio-metabolic health outcomes, and whether these differ by offspring age.

**Design:** Multi-cohort study.

**Setting:** Fourteen population-based cohort studies with offspring from the UK, Ireland, France, the Netherlands, Portugal, Greece, Italy, Norway, Singapore, and Australia for meta-analysis of various ages. Four cohorts (three European and one Singaporean) with repeated measures for pooled age-change (from 3 to 26 years) trajectory analysis.

**Participants:** Young people sampled from the general population with complete data on mode of conception, confounders, and ≥1 cardio-metabolic outcome measured after birth.

**Exposures:** Conception by ART versus natural conception (NC).

**Main outcome measures:** Systolic (SBP) and diastolic blood pressure (DBP), heart rate (HR), total cholesterol (TC), high-density lipoprotein cholesterol (HDLc), low-density lipoprotein cholesterol (LDLc), triglycerides (TG), glucose, insulin, and glycated haemoglobin (HbA1c).

**Results:** Between 35,780 (605 ART) and 4,502 (67 ART) offspring were included in meta-analysis of various ages for each outcome. Mean age at outcome assessment ranged from 13 months to 27.4 years, with most cohorts ((11/14) having mean age <10 years. Compared with NC, ART-conceived offspring had similar SBP (mean difference (ART minus NC): -0.89*mmHg*; 95%CI: -1.91 to 0.14), DBP (−0.50*mmHg*; -1.65 to 0.66), and HR (0.02*beats/min*; -1.00 to 1.03). Cholesterol measures were higher in ART-conceived than NC offspring, for TC (mean % difference: 2.54*%*; 0.46 to 4.61), HDLc (4.17*%*; 1.79 to 6.56), and LDLc (4.95*%*; 0.99 to 8.92), whereas triglycerides were similar (−1.53*%*; -6.19 to 3.13). No clear differences were seen for glucose (0.25*%*; -1.38 to 1.88), insulin (−5.04*%*; -13.20 to 3.12), or HbA1c (−0.07*%*; -0.14 to 0.00). Trajectory models in up to 17,244 (244 ART) offspring showed that early life trajectory differences were consistent with the above pooled results and showed higher SBP emerging from mid-adolescence to adulthood with ART (e.g., predicted mean difference in SBP at age 26 years for ART versus NC was 5.06*mmHg*; 1.76 to 8.35).

**Conclusions:** Children conceived through ART had higher cholesterol and similar blood pressure and hyperglycaemic/insulin resistance measures compared with NC children. Whilst overall this is reassuring, our trajectory analysis in a sub-group of cohorts suggested that those conceived by ART may go on to develop higher blood pressure in early adulthood. Our study shows the importance of follow-up into adulthood and requires validation by independent studies with different study designs including within-sibship and mechanistic studies.

## INTRODUCTION

Use of assisted reproductive technology (ART), which mainly involves *in vitro* fertilisation (IVF) or intracytoplasmic sperm injection (ICSI), has risen rapidly in developed countries in recent decades leading to >8 million births worldwide, and this is expected to continue to rise (1). There is concern that use of ART may cause adverse cardiovascular and metabolic health outcomes in the offspring (2). Systematic reviews of what are mostly small studies report that ART conception is associated with higher offspring blood pressure, glucose, and triglycerides (3, 4) however, publication and selection bias might influence these findings. Selection bias could arise as most previous studies were clinical cohorts of ART conceptions compared to select naturally conceived (NC) comparison groups (e.g., family friends) who were not followed-up from conception in the same way as those conceived by ART.

A Swiss study published since these reviews that included 54 ART-conceived and 43 NC children discovered signs of premature vascular ageing which persisted at 5-year follow-up assessments at age 17 years, along with new evidence of higher blood pressure that emerged at this older age (5). However, family friends were used as NC controls which may introduce a selection bias. A more recent Singaporean population-based birth cohort study where both ART-conceived and NC offspring were selected form the same underlying population and followed-up in the same way (N=1,180 with 85 ART-conceived offspring) found that ART-conceived offspring had lower blood pressure from age 3 to 6 years (6). To the best of our knowledge, no large population-based studies of cardio-metabolic health outcomes in ART-conceived offspring, or studies that explored how associations change with increasing age, are available. It is important to explore how associations evolve with age as we cannot assume that associations in early childhood will persist through adulthood.

Our aim was to conduct a large population-based multi-cohort study with longitudinal repeated measurements analysis to provide more reliable evidence (and so also limiting potential publication bias) on associations of ART conception with long-term offspring cardio-metabolic health up to young adulthood. Additionally, we investigated the role of underlying parental subfertility, compared associations according to sex and types of ART, and explored if results were driven by multiple births, prematurity, and offspring adiposity.

## METHODS

This study was carried out by following a pre-specified analysis plan and code developed by AE, AT, HI, and DAL (https://osf.io/qhwvc/) and is reported in line with The Strengthening the Reporting of Observational Studies in Epidemiology (STROBE) Statement (7).

### Cohort studies

Cohort studies were recruited from the Assisted Reproductive Technology and future Health (ART-Health) cohort collaboration (8). Briefly, ART-Health is a collaboration between 26 multinational cohort studies that were recruited from the EU Child Cohort Network and elsewhere (9, 10) with specific targeting of population-based studies without selection or oversampling of those conceived by ART to avoid a selection bias and ensure identical outcome assessments for ART-conceived and NC offspring. Cohorts were eligible to be included in the current analysis in they had data on whether offspring were conceived by ART or not, and ≥1 cardio-metabolic health measure assessed at any age after birth. In total, 14/26 cohorts had relevant data and were included in this study (**Supplementary Text 1**).

All included cohorts had approval from their relevant local/national ethics committees and study participants gave informed consent/assent to participate in the respective cohorts and secondary data analyses. Detail on ethics approvals/consent in each cohort can be found in **Supplementary Text 1**.

### Mode of conception

Data on mode of conception and fertility treatment (11) was gathered from record linkage or questionnaires (**Supplementary Text 1**). For our main analysis, a dichotomous variable was used to compare offspring conceived by ART (IVF or ICSI) with NC offspring. Where data were available, we considered ART treatment type (IVF vs ICSI), whether the offspring were conceived by fresh embryo transfer (fresh ET) or frozen embryo transfer (FET), and whether parents of NC offspring were fertile or sub-fertile, depending on length of time to pregnancy being ≤12 months or >12 months since started trying, respectively.

### Offspring cardio-metabolic outcomes

Eligible offspring cardio-metabolic outcomes were systolic blood pressure (SBP, mmHg), diastolic blood pressure (DBP, mmHg), heart rate (HR, beats per minute), total cholesterol (TC, mmol/l), high-density lipoprotein cholesterol (HDLc, mmol/l), low-density lipoprotein cholesterol (LDLc, mmol/l), triglycerides (TG, mmol/l), glucose (mmol/l), insulin (mu/l) and glycated haemoglobin (HbA1c, %). Similar protocols were followed across cohorts, and full details on outcome measurements are in **Supplementary Text 1**. Briefly, SBP, DBP, and HR were measured using blood pressure monitors with participants seated and at rest, with SBP, DBP and HR calculated as the average of first, second, and (if available) third measurements. Biomarkers (lipids, hyperglycaemic/insulin resistance markers) were obtained using standard clinical laboratory procedures in fasting or non-fasting blood samples, depending on age. To maximise sample size for the meta-analysis of all cohorts, if a cohort had repeated measures of an outcome, we selected the age with the largest number of offspring (**Supplementary Table 1**). For the age-change trajectory analysis, all repeated measurements (and all cohorts that had repeated measures and were able to share their data) were included in the analysis.

### Confounders

We used Directed Acyclic Graphs, developed with input from the multidisciplinary author group, to identify and control for confounders, and to avoid over-adjustment for mediators (**Supplementary Figure 1**). This identified the following potential confounders: maternal age at pregnancy/birth, parity, pre-pregnancy BMI and smoking, education (as a marker of socioeconomic position) and ethnicity. Most (12/14) cohorts had data on all confounders; two (HUNT and CHART) were unable to adjust for maternal BMI, smoking or ethnicity, with one of them also unable to adjust for maternal education. Details on how confounders were measured in each cohort is in **Supplementary Text 1**.

### Statistical analysis

#### Analysis of all cohorts (various ages)

Associations of ART conception with cardio-metabolic outcomes were examined separately in each cohort and results were subsequently combined using meta-analysis. Cohort-specific linear regression models were used to estimate mean difference and 95% confidence intervals (CIs) for each cardio-metabolic outcome between ART-conceived and NC offspring. Models were adjusted for confounders (maternal age, education, parity, BMI, smoking, ethnicity) and offspring sex and age (to aid precision), and robust standard errors were used by cohorts with related individuals (i.e., siblings, twins, or cousins). To allow comparison of results between outcomes, SBP, DBP, and HR were analysed in cohort-specific standard deviation (SD) units with mean=0 and SD=1 (with analyses in original units also performed). Lipids, glucose, and insulin were analysed after natural log transformation and the results presented as percentage (%) differences between ART-conceived and NC offspring (12), This was done because these biomarkers, and hence the regression model residuals, were right skewed, and to facilitate comparability of the results across markers. HbA1c was analysed in its original units (%).

Cohort results were pooled using random-effects meta-analysis to obtain mean differences and 95% CIs in outcomes across all included cohorts. *I*^*2*^ was used to quantify the consistency in these results as the percentage of total variability due to between-cohort heterogeneity (13). Robustness of the pooled results to influential cohorts was investigated using a leave-one-out analysis where the meta-analysis model was repeated by leaving one of the cohorts out each time (14). The contribution of each cohort to overall heterogeneity and the impact on pooled estimates were graphically represented in a modified Baujat plot (14). Meta-analysis was also repeated after excluding two cohorts that were unable to adjust for maternal BMI, smoking and ethnicity.

The following prespecified sub-group analyses were performed to explore sources of heterogeneity. We explored whether results differed by offspring mean age at outcome assessment in sub-group meta-analyses that compared results across cohorts with mean age <10 years versus 10 or older. We attempted to separate out effects of ART conception from effects of parental subfertility, by repeating analyses comparing ART-conceived with NC offspring from sub-fertile parents and to offspring from fertile parents. Differences by sex and ART treatment were explored by repeating analyses stratified by sex; comparing IVF and ICSI separately to NC; and comparing fresh ET and FET separately to NC and inspecting the difference in effect sizes between groups. Lastly, we explored whether results were driven by twins/multiple births by repeating analyses in singletons and examined if results were driven by prematurity and offspring adiposity by refitting the main confounder-adjusted models with extra adjustment for offspring birth weight, gestational age at delivery and BMI (before or at outcome assessment).

#### Age-change (3 to 26 years) trajectory analysis

Differences in cardiovascular (SBP, DBP, HR), lipids (TC, HDLc, LDLc, TG), and glucose trajectories from childhood to young adulthood between ART-conceived and NC offspring were examined in four cohort studies that collected repeated measurements: (i) the UK-based Avon Longitudinal Study of Parents and Children (ALSPAC) with between 1 to 12 repeated blood pressure and heart rate measures, 1 to 7 repeated lipids measurements (from age 3 to 26 years), and 1 to 4 repeated glucose measurements from age 7 to 26 years (15-17), (ii) the Portuguese G21 cohort with 1-3 repeated measurements taken at ages 4, 7, and 10 years for all outcomes (18), (iii) Amsterdam Born Children and their Development study (ABCD) with up to 2 repeated measures for all outcomes at ages 5 and 11 years (19), and (iv) the Growing up in Singapore Towards healthy Outcomes (GUSTO) study (20) with 1 to 6 repeated SBP, DBP, and HR measurements from age 3 to 8 years, and 2 lipids and glucose measurements at ages 6 and 8 years.

Trajectories were analysed using natural cubic spline mixed effects models (21), which were adjusted for sex, confounders, and cohort, and included an interaction term (age by ART) to allow different cardio-metabolic trajectories for ART-conceived and NC offspring (21, 22). Predicted mean trajectories and differences were calculated. We explored whether trajectory differences reflected multiple births by repeating models in singletons and explored whether differences were driven by prematurity and offspring adiposity by refitting the confounder-adjusted models with extra adjustment for birth weight, gestational age, and offspring BMI-for-age Z score (taken before the first outcome measurement and standardized to the WHO Growth reference standards). Analysis was done in R version 4.0.2 (R Project for Statistical Computing) (23-25).

#### Missing data

For the analyses of all cohorts (with various ages), all offspring were included if they had data on mode of conception, confounders, and the specific cardio-metabolic outcome. In the age-change trajectory analyses, we included in each analysis all offspring with data on mode of conception, confounders and at least one outcome measure under the missing at random assumption. To explore the potential impact of missing data we compared baseline maternal characteristics between included offspring and those that were excluded due to missing data (**Supplementary Table 2**).

### Patient and public involvement

This research did not involve patients or the public as it uses data that were previously obtained from cohorts of people who had already been recruited. As such, no patients or member of the public were involved in the design or implementation of this study. However, the long-tern health of ART-conceived children has been identified as a research priority by the parents of those conceived through ART (26).

## RESULTS

### Participant characteristics

A total of 14 mostly European cohort studies were included (two cohorts each from the UK, the Netherlands, Italy, and Australia, and one cohort each from Ireland, France, Portugal, Greece, Norway, and Singapore). Birth years ranged from 1984 to 2018, though most were born from 2002 onwards. Mean age at each outcome assessment ranged from 13 months to 27.4 years, but most (in 11/14 cohorts) had mean age <10 years. The number of cohorts and offspring included in the main meta-analysis (i.e., ART compared with NC) ranged from 14 cohorts and 35,780 (605 ART) offspring for SBP to 2 cohorts and 4,502 (67 ART) offspring for HbA1c. The number of offspring from each cohort in each analysis and the mean and SD of the outcomes and ages at each outcome assessment are given in **Supplementary Table 1**. Those excluded due to missing data had lower maternal education and higher prevalence of pregnancy smoking but were broadly similar on the other maternal factors (**Supplementary Table 2)**

### Results of analysis with all cohorts/ages

The pooled confounder-adjusted mean differences in cardio-metabolic outcomes between ART-conceived and NC offspring are presented in **Figure 1**, with results from each cohort presented, arranged by mean age, in **Supplementary Figure 2**. SBP (mean difference between ART-conceived and NC across all cohorts: -0.09SD; 95%CI: -0.21 to 0.03) and DBP (−0.07SD; -0.22 to 0.08) were slightly lower in ART-conceived than NC offspring, whereas mean HR (0.00SD; -0.09 to 0.10) was similar in both groups. Mean TC (mean % differences: 2.54%; 0.46 to 4.61), HDLc (4.17%; 1.79 to 6.56), and LDLc (4.95%; 0.99 to 8.92) were all higher in ART-conceived than NC offspring, whereas mean TG (−1.53%; -6.19 to 3.13) was similar in both groups. No clear difference was seen for glucose (mean % differences: 0.25%; -1.38 to 1.88), with some evidence of slightly lower insulin (−5.04%; -13.20 to 3.12) and HbA1c (−0.07%; -0.14 to 0.00) in ART-conceived offspring (**Figure 1**).

**Figure 1.**
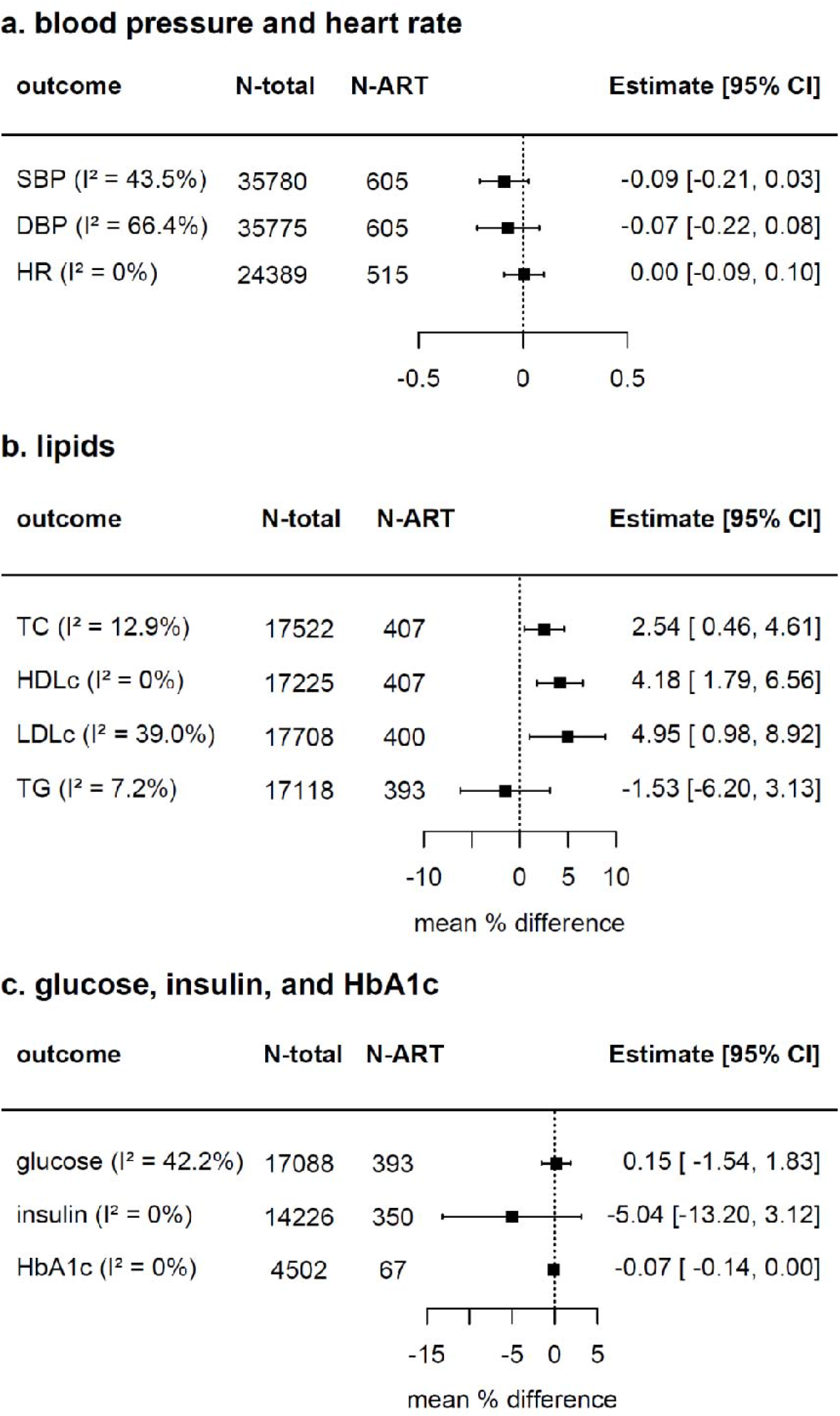
Pooled mean differences in cardio-metabolic health outcomes between ART-conceived and NC offspring from up to fourteen birth cohort studies Figure shows the pooled confounder-adjusted mean differences in SD units [and 95% confidence intervals] in cardio-metabolic outcomes between ART-conceived and NC

There was no evidence of heterogeneity between cohorts for HR, HDLc, insulin, HbA1c (*I*^*2*^ =0% for all) and TG (*I*^*2*^ =7.2%) (**Supplementary Figure 3**). Evidence of heterogeneity was found for SBP (*I*^*2*^=43.4%), DBP (*I*^*2*^ =66.1%), LDLc (*I*^*2*^ =39.0%), and glucose (*I*^*2*^=42.2%) results. Influence diagnostics showed that the HUNT cohort (one of the two cohorts with missing confounders) had a major influence on the pooled results for DBP (**Figure 2**); the pooled estimate was attenuated when HUNT was removed (mean difference in DBP between ART-conceived and NC across all cohorts with HUNT excluded was -0.03SD; -0.12 to 0.06). LDLc results were influenced by the G21 and Generation R cohorts; removing G21 pulled results towards higher LDLc with ART whereas removing Generation R pulled results to a smaller positive association (**Figure 2**). G21 had a considerable impact on glucose results, removing G21 pulled estimate to higher glucose in ART-conceived offspring (**Figure 2**).

**Figure 2.**
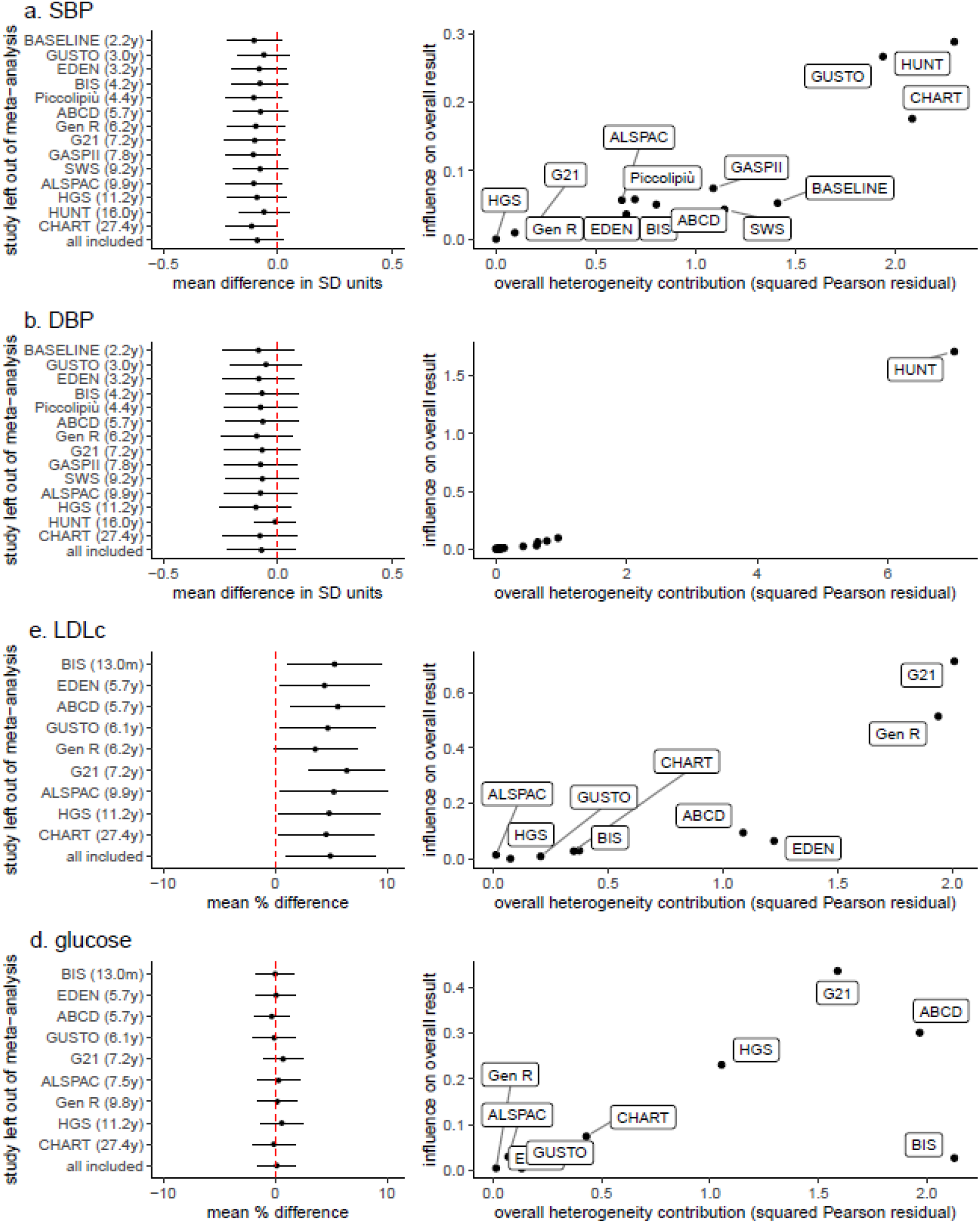
Results from leave-one-out diagnostics analysis shown for outcomes where there was some evidence of inconsistency across cohorts offspring from up to 14 cohort studies. Cohort specific models were adjusted (as fully as possible) for maternal age, parity, BMI, smoking, education, and ethnicity, plus offspring age and sex. The results from each cohort are presented in **Supplementary Figure 2**. ART: assisted reproductive technology. NC: natural conception. SBP: systolic blood pressure, DBP: diastolic blood pressure, HR: heart rate, TC: total cholesterol, HDLc: high-density lipoprotein cholesterol, LDLc: low-density lipoprotein cholesterol, TG: triglycerides, HbA1c: glycated haemoglobin. The left panels show the pooled confounder-adjusted estimates from random-effects meta-analysis when leaving each cohort out in turn, as well the estimate for all cohorts combined (i.e., the results shown in **Figure 1**). The right panels show modified Baujat plots where the x-axis corresponds to the squared Pearson residual of each cohort, and y-axis corresponds to the standardized squared difference between the fitted value for the cohort, with and without the cohort included in the meta-analysis model. Results are presented for systolic (SBP) and diastolic blood pressure (DBP), low-density lipoprotein cholesterol (LDLc), and glucose. Results for all cardio-metabolic outcomes are shown in **Supplementary Figure 3**.

There was no evidence from sub-group meta-analysis that results for blood pressure, HR, HDLc, and glucose differed between offspring aged under 10 years versus older offspring, including in models without the two cohorts that were unable to adjust for maternal BMI, smoking and ethnicity (**Supplementary Table 3**). There was imprecise evidence that TC, and LDLc were higher for ART-conceived (versus NC offspring) aged <10 years but not for older offspring, and that insulin was lower in ART-conceived offspring aged 10 year or older than NC offspring (**Supplementary Table 3**).

Results were similar when ART-conceived offspring were compared with NC offspring from sub-fertile and fertile parents, and when IVF and ICSI were each compared with NC (**Figure 3**). Mean blood pressure was lower for ART-conceived than NC offspring in males but not in females, and for offspring conceived by fresh ET versus NC but was similar for FET versus NC (**Figure 3**). Mean LDLc was higher in ART-conceived compared with NC offspring in females but not males (**Figure 3**). Lipids were similar in fresh ET and NC groups but were higher in offspring conceived from FET versus NC (**Figure 3**). Insulin was lower in offspring conceived by fresh ET than in NC offspring and was slightly higher for FET compared with NC though confidence intervals were wide (**Figure 3**).

**Figure 3.**
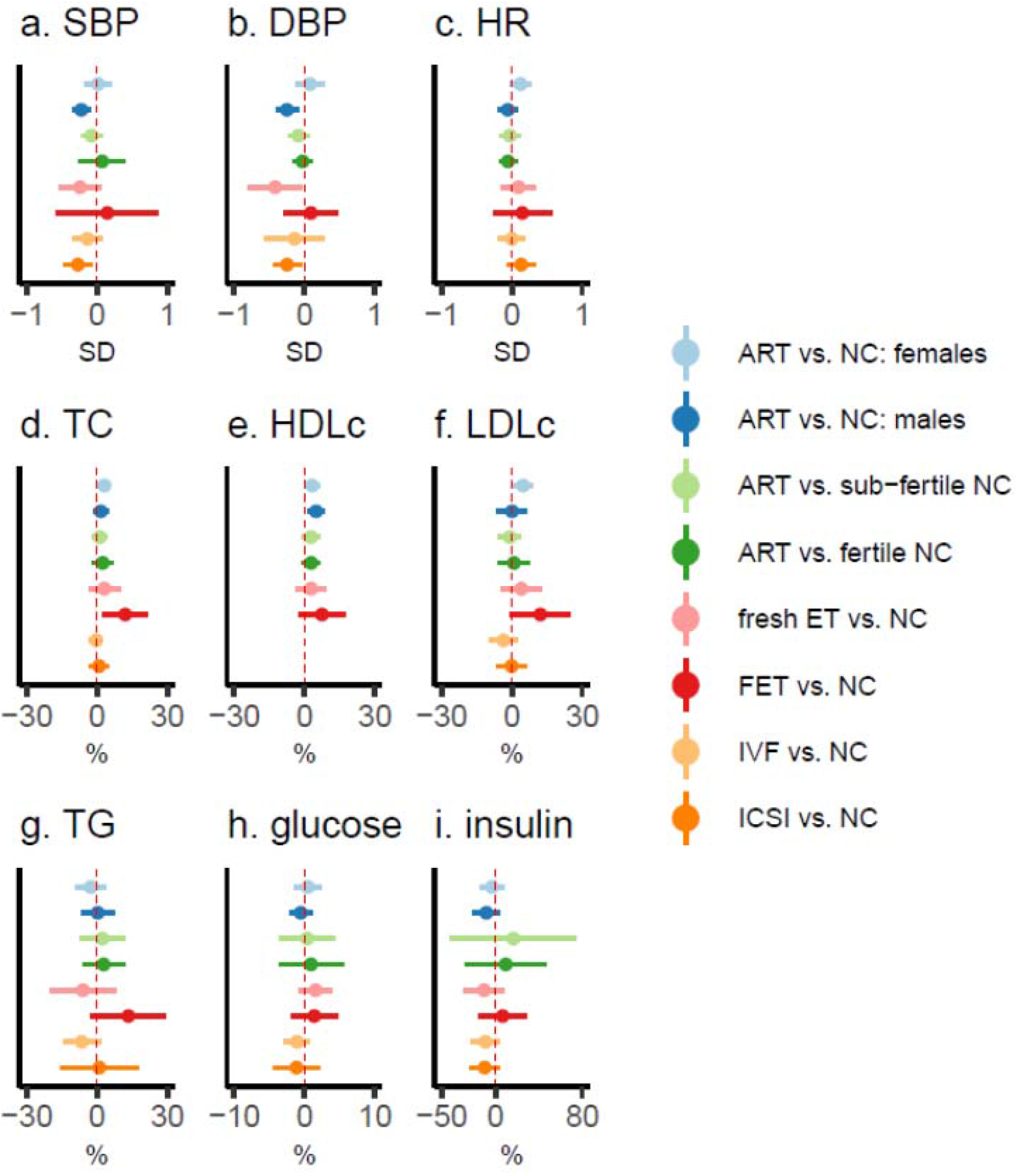
Pooled mean differences in cardio-metabolic outcomes between ART-conceived and NC offspring, stratified by sex, parental sub-fertility, fresh/frozen embryo transfer, and IVF/ICSI Figure show pooled confounder-adjusted mean differences in SD units [and 95% confidence intervals] in cardio-metabolic outcomes between ART-conceived and NC offspring. Cohort specific models were adjusted (as fully as possible) for maternal age, parity, BMI, smoking, education, and ethnicity, plus offspring age and sex. ART: assisted reproductive technology. NC: natural conception. SBP: systolic blood pressure, DBP: diastolic blood pressure, HR: heart rate, TC: total cholesterol, HDLc: high-density lipoprotein cholesterol, LDLc: low-density lipoprotein cholesterol, TG: triglycerides, HbA1c: glycated haemoglobin. IVF: in vitro fertilisation, ICSI: intracytoplasmic sperm injection, ET: embryo transfer. FET: frozen embryo transfer.

Results in singleton births were consistent with results in all participants including those from multiple births (**Supplementary Figure 4**), and differences in blood pressure were increased by adjustment for birth weight, gestational age and offspring BMI, and were similar to the confounder-adjusted results for all other outcomes (**Supplementary Figure 5)**.

### Results of age-change trajectory analysis

A total of 17,244 (244 ART), 16,818 (243 ART), 13,9126 (188 ART), and 13,386 (184 ART) offspring were included in pooled trajectory analysis for blood pressure, heart rate, lipids, and glucose, respectively. The predicted mean trajectories for ART-conceived and NC offspring are shown in **Figure 4** and the predicted mean differences in each cardio-metabolic outcome from childhood to adulthood are presented in **Figure 5** and **Supplementary Table 4**.

**Figure 4.**
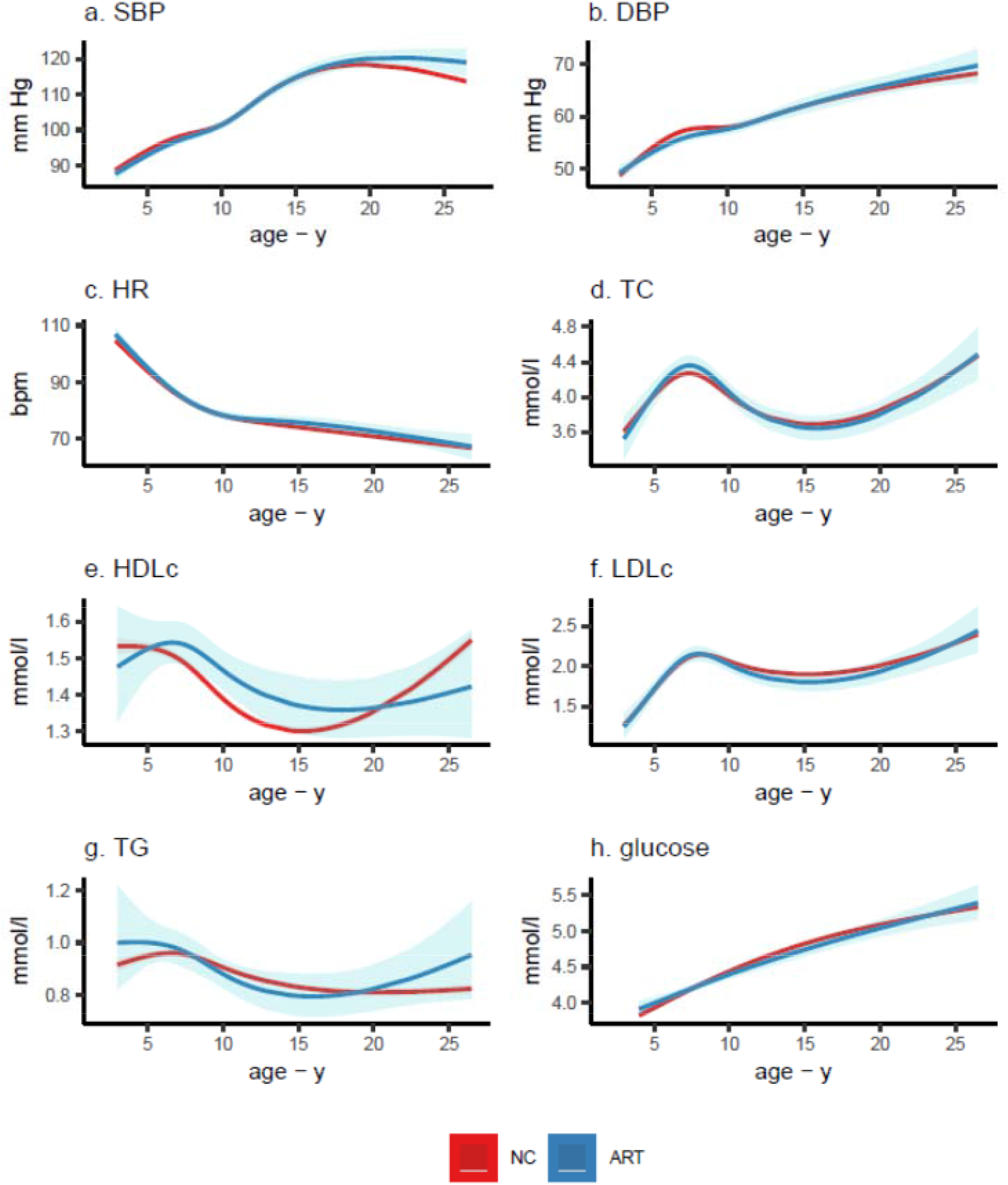
Predicted mean cardio-metabolic trajectories from age 3-26 years in ART-conceived and NC offspring from four population-based birth cohort studies Figure shows the predicted mean trajectories in cardio-metabolic outcomes from childhood to adulthood for ART-conceived and NC offspring. Predicted means were obtained from multicohort (ALSPAC, G21, ABCD, and GUSTO cohorts) natural cubic spline mixed effects trajectory models. Models were adjusted for offspring sex, maternal age, parity, BMI, smoking, education, and ethnicity, and included an interaction between ART and age. SBP: systolic blood pressure, DBP: diastolic blood pressure, HR: heart rate, TC: total cholesterol, HDLc: high-density lipoprotein cholesterol, LDLc: low-density lipoprotein cholesterol, TG: triglycerides. All models included 2 knots placed at quantiles of age, except for glucose models which included 1 knot placed at median age. A total of 17,244 (244 ART), 16,818 (243 ART), 13,9126 (188 ART), and 13,386 (184 ART) offspring were included in the analyses of systolic/diastolic blood pressure, heart rate, lipids, and glucose trajectories, respectively.

**Figure 5.**
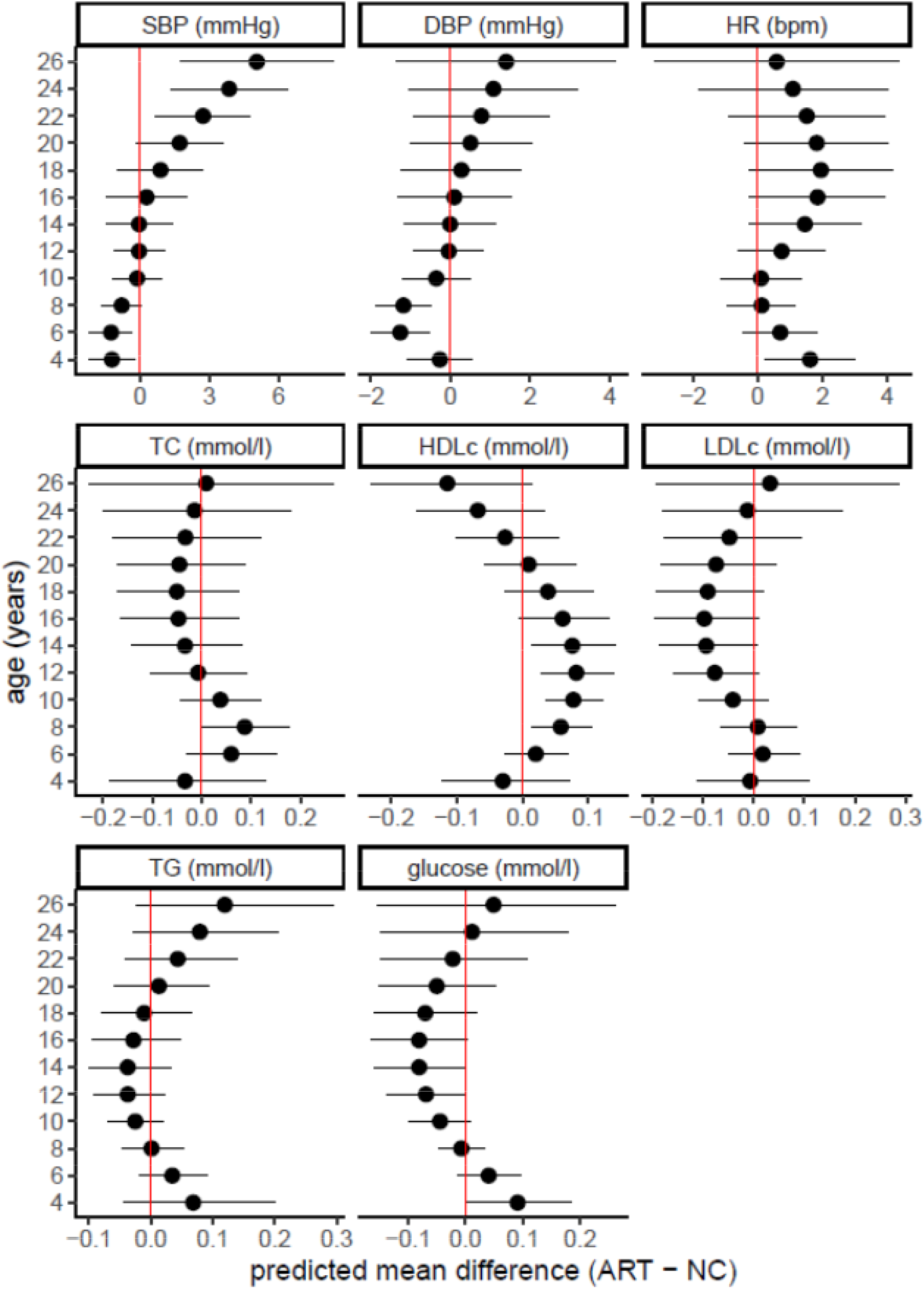
Predicted mean differences in cardio-metabolic outcomes from childhood to adulthood between ART-conceived and NC offspring Figure shows predicted mean differences (and 95%CIs) in cardio-metabolic outcomes from childhood to adulthood between ART-conceived and NC offspring. Predicted means were obtained from multicohort (ALSPAC, G21, ABCD, and GUSTO cohorts) natural cubic spline mixed effects trajectory models, that were adjusted for offspring sex, maternal age, parity, BMI, smoking, education, and ethnicity. All models included an interaction between ART and age. SBP: systolic blood pressure, DBP: diastolic blood pressure, HR: heart rate, TC: total cholesterol, HDLc: high-density lipoprotein cholesterol, LDLc: low-density lipoprotein cholesterol, TG: triglycerides

Blood pressure was slighter lower during childhood in ART-conceived offspring (e.g., the predicted differences (ART minus NC) at age 6 years were -1.26mmHg (−2.19 to -0.34) for SBP and -1.26mmHg (−1.99 to -0.54) for DBP). Blood pressure was then more similar with older age in both groups until age 14-16 years. Subsequently, SBP appeared higher in ART-conceived offspring, with similar but less marked change in DBP. For example, the predicted differences at age 26 years were 5.06mmHg (1.76 to 8.35) for SBP and 1.40mmHg (−1.36 to 4.15) for DBP. HR appeared slightly higher in ART offspring throughout follow-up, but the difference was small, e.g., predicted differences at age 4 and 26 years were 1.61bpm (0.22 to 3.01) and 0.58bpm (−3.19 to 4.35), respectively.

There was no clear evidence of difference in TC at all ages except for suggestion of higher levels at age 8 for ART-conceived offspring (predicted difference in TC: 0.09mmol/l (0.00 to 0.18). HDLc appeared higher in ART-conceived offspring from childhood to adolescence and then appeared slightly lower in adulthood, e.g., predicted differences in HDLc at age 14 and 26 years were 0.08mmol/l (0.01 to 0.14) and -0.11mmol/l (−0.23 to 0.01), respectively. There was some suggestion for higher TG in early childhood, similar levels during adolescence and higher levels in adulthood but differences were imprecise across all ages, e.g., predicted differences in TG at ages 4, 16 and 26 years were 0.07mmol/l (−0.04 to 0.2), -0.03mmol/l (−0.1 to 0.05), and 0.12mmol/l (−0.02 to 0.29), respectively. No noticeable differences were seen in LDLc or glucose trajectories.

Lastly, predicted mean trajectories from analyses in singletons (**Supplementary Figure 6**), and with extra adjustment for birth weight, gestational age (**Supplementary Figure 7**), and offspring BMI (**Supplementary Figure 8**) were consistent with results of the confounder-adjusted models.

## DISCUSSION

### Principal findings

Our multi-cohort study with pooled results from more than 35,000 mostly young children aged under 10 years found no clear adverse differences in triglycerides, blood pressure, heart rate or hyperglycaemic/insulin resistance traits but some evidence for a higher cholesterol in ART-conceived compared with NC children. Complementary follow-up analyses of cardio-metabolic health trajectories from age 3 to 26 years in more than 17,000 (244 ART) offspring identified adverse mean trajectories in blood pressure from early adolescence, with the differences increasing with older age; this resulted in mean SBP becoming higher in adult ART-conceived offspring. Results were consistent when comparing ART-conceived offspring with NC offspring with/without parental subfertility, fresh ET and FET with NC, and conventional IVF and ICSI with NC, as well as when restricted to singletons, and with adjustment for birth weight, gestational age and own BMI.

### Comparison with other studies

To the best of our knowledge, this is the largest study focusing upon cardiovascular and cardio-metabolic outcomes in ART-conceived offspring, and the longest follow-up of its kind. Our finding of slightly lower SBP across all cohorts, which our trajectory analysis showed, was only seen up to age 8 years, is consistent with recent evidence from a small Singaporean birth cohort showing lower SBP across four time-points from age 3 to 6 years (6). Our finding that this result switches to higher blood pressure from early adolescence is consistent with findings from a small Swiss clinical ART cohort showing that higher blood pressure was only observed after 5-year follow-up to age 17, despite evidence of premature vascular ageing observed at both baseline and follow-up (5). Our novel trajectory results add evidence that higher SBP persists up to age 26 years, in addition to suggestive evidence for higher DBP and TG observed at this age.

While the underlying mechanisms by which ART can lead to differences in cardio-metabolic health are unknown and require investigation, one explanation for the childhood to adulthood patterns of differences in blood pressure, as well as possibly adverse TG trajectories at older ages, is that they reflect effects of age-specific adiposity differences. This is supported by our recent findings from 26 ART-Health cohorts showing smaller size and lower adiposity during early life in ART-conceived than NC offspring and a subsequently higher adiposity in young adulthood (8), which may in turn influence cardio-metabolic traits (27). This higher adiposity in young adulthood in ART-conceived offspring was also found in a recent analysis of young adults from Nordic registries (28). If validated, our current findings in this study suggest that while adolescent or young adult conceived by ART are too young to have evidence of clinical metabolic or cardiovascular diseases in, underlying pre-clinical risk factors could exist (5). These factors (higher blood pressure and TG) track across life and have been shown to increase cardiovascular disease risk in into later life (29-32).

### Strengths and limitations of the study

Strengths of this study include the large sample size in comparison with previous studies and the inclusion of cohorts from a range of geographic regions, which should make the findings generalisable to more populations. The use of cohorts with comparison groups from the same underlying population as those conceived by ART is another important strength, which is not the case with many clinical cohorts where controls are selected from relatives or friends of the couples undergoing ART. Ou novel trajectory analysis is another strength as it allows both change with age and, importantly, a wider age range to be explored, thus giving insight into evolution of cardio-metabolic risk factors over the life course.

Study limitations include the low precision/power for some outcomes even with this large collaboration, and most cohorts being young meant we were not able to assess results by age groups and we could only do trajectory analyses in a sub-group of the included cohorts. Our analysis of all cohorts was restricted to offspring with complete data on mode of conception confounders, and outcome which may have reduced precision of estimates and introduced a bias due to missing data. Our trajectory analysis approach allowed the inclusion of all those with at least one outcome measure under the missing at random assumption, which reduces bias due to missing outcome data however, those missing exposure and confounders were excluded from this analysis. Our analytical samples were socioeconomically different from those excluded due to missing data, which might limit the generalisability of our findings. Residual confounding from unmeasured confounders such as parental cardio-metabolic health and paternal factors is possible and may influence our findings.

### Conclusion and implications

In summary, our study found evidence of raised lipids in ART-conceived compared with NC children, and that ART-conceived offspring had more adverse blood pressure trajectories and higher blood pressure from adolescence to young adulthood, with no adverse differences in heart rate or glucose-related traits from childhood up to young adulthood. While our results require validation by independent studies with different study designs like within-sibship and mechanistic studies, they indicate that ART-conceived offspring might therefore benefit from indicated cardiometabolic screening, Future research on metabolomics and cardiovascular or arterial phenotypes may provide insight into possible underlying mechanisms.

## Data Availability

Data used in this study is available to bone fide researchers upon request to each cohort. Details of how to access the data is provided in the Supplement.

## DECLARTIONS

### Contributors

DAL conceived the study. DAL and AE wrote the first draft of the analysis plan with input from AT and HI. AE did analysis in ALSPAC and ABCD. TS did analysis in SWS. GMM did analysis in BASELINE. MT did analysis in EDEN. CR did analysis in Gen XXI. GSE did analysis in Gen R. FA did analysis in GASPII. LG did analysis in Piccolipiù. MCM did analysis in MoBa and HUNT. JH did analysis in GUSTO. TM did analysis in BIS and CHART. AE did the meta-analysis and trajectory analysis. DAL and AE wrote the first manuscript draft and all authors provided comments and approved the final version.

### Funding

This project has received funding from the European Research Council (ERC) under the European Union’s Horizon 2020 research and innovation programme grant agreement No. 101021566 (ART-HEALTH) and No. 733206 (LifeCycle), the UK Medical Research Council (MC_UU_00011/6), the British Heart Foundation (CH/F/20/90003 and AA/18/7/34219) and the Bristol National Institute of Health Research Biomedical Research Centre. Cohort and author specific funding is provided in **Supplementary Text 2**.

### Ethical approval

All cohorts included in this study received ethical approvals from their respective ethical advisory boards and all obtained informed participant/parental consent. Details on ethical approval and consent for each cohort is in **Supplementary Text 1**.

### Data sharing

Data used in this study is available to bone fide researchers upon request to each cohort. Details of how to access the data is provided in **Supplementary Text 2**. Please contact Professor Deborah Lawlor and Dr Ahmed Elhakeem if you have relevant data and would like to join the ART-HEALTH consortium and contribute to future collaborations.

