## Supplementary material for "Association of assisted reproductive technology with long-term offspring cardiometabolic health: a multi-cohort study"

**Association of assisted reproductive technology with offspring cardio-metabolic health: a multi-cohort study (supplementary material)**

Ahmed Elhakeem^1,2^, Amy E Taylor^1,2,3^, Hazel M Inskip^4^, Jonathan Huang^5,6^, Toby Mansell^7,8^, Carina Rodrigues^9,10^, Federica Asta^11^, Sophie M Blaauwendraad^12,13^, Siri E Håberg^14^, Jane Halliday^7,8^, Margreet W Harskamp-van Ginkel^15^, Jian-Rong He^16^, Vincent WV Jaddoe^13,12^, Sharon Lewis^7,8^, Gillian M Maher^17,18^, Yannis Manios^19,20^, Fergus P McCarthy^21,18^, Irwin KM Reiss^13^, Franca Rusconi^22^, Theodosia Salika^4^, Muriel Tafflet^23^, Xiu Qiu^16^, Bjørn O Åsvold^24,25,26^, David Burgner^7,27,28^, Jerry KY Chan^29,30^, Luigi Gagliardi^22^, Romy Gaillard^13,12^, Barbara Heude^23^, Maria C Magnus^14^, George Moschonis^31^, Deirdre Murray^32,18^, Scott M Nelson^3,33^, Daniela Porta^11^, Richard Saffery^7,8^, Henrique Barros^9,10^, Johan G Eriksson^5,34,35,36^, Tanja GM Vrijkotte^15^, Deborah A Lawlor^1,2,3^

### Ethics approvals

| Cohort name | Ethic approval description |
| --- | --- |
| ABCD | Approval for the ABCD study was obtained from the Central Committee on Research involving Human Subjects in the Netherlands, the Medical Ethical Committees of the participating hospitals, and from the Registration Committee of the Municipality of Amsterdam. Written informed consent was obtained from all participating mothers. |
| ALSPAC | Ethical approval for the study was obtained from the ALSPAC Ethics and Law Committee and the Local Research Ethics Committees. Informed consent for the use of data collected via questionnaires and clinics was obtained from participants following the recommendations of the ALSPAC Ethics and Law Committee at the time. At age 18, study children were sent 'fair processing' materials describing ALSPAC’s intended use of their health and administrative records and were given clear means to consent or object via a written form. Data were not extracted for participants who objected, or who were not sent fair processing materials. Ethical approval for the study was obtained from the ALSPAC Law and Ethics committee and local research ethics committees (NHS Haydock REC: 10/H1010/70). |
| BASELINE | Research objectives and measurements in this birth cohort were conducted according to the guidelines laid down in the Declaration of Helsinki and all procedures were approved by the Clinical Research Ethics Committee of the Cork Teaching Hospitals, [ref ECM5(9) 01/07/ 2008]. Families provided written informed consent at 20 weeks’ gestation or at birth to participate in BASELINE follow-up. |
| BIS | Ethics approval was obtained from the Barwon Health Human Research Ethics Committee (10/24). All mothers provided written informed consent. |
| CHART | The study was approved by the Royal Children's Hospital Human Research Ethics Committee, and all study participants provided consent to take part in the study. |
| EDEN | The study received approval from the ethics committee (CCPPRB) of Kremlin Bicêtre on 12 December 2002 and from CNIL (Commission Nationale Informatique et Liberté), the French data privacy institution. All subjects gave their informed consent for inclusion before they participated in the study. Consent for the child was obtained from both parents after the child's birth. |
| GASPII | The protocol of the study has been approved by the Ethics committees of the Università Cattolica del Sacro Cuore, Rome, and all study participants provided consent to take part in the study. |
| Gen-R | The study has been approved by the Medical Ethical Committee of the Erasmus MC, University Medical Center in Rotterdam (MEC-2012-165-NL40020.078.12). Written informed consent was obtained from the parents or legal representatives of the children. Even with consent of the parents, when the child is not willing to participate actively, no measurements are performed. |
| G21 | The Ethics Committee of Hospital de São João, and of the Institute of Public Health of the University of Porto approved the study protocols. The study complies with the Ethical Principles expressed in the Helsinki Declaration and with the national legislation and was registered with the Portuguese Authority for Data Protection. In all evaluations, participants were informed about the purposes and design of the study, as well as the potential discomfort caused by participation. Signed informed consent was obtained from all parents or legal guardians, and oral assent was obtained from children at each evaluation. |
| GUSTO | Study protocols following the principles of the Declaration of Helsinki and were approved by the respective ethics committees for two hospitals: National Healthcare Group Domain Specific Review Board (NUH) and SingHealth Centralized Institutional Review Board (KKH). All participants in this study provided informed consent to participate and contribute their data to publications. The GUSTO study is registered under study ID: NCT01174875 (clinicaltrials.gov) which broadly covers investigations of parental and gestational influences on child health. |
| HGS | Approval to conduct the study was granted by the Greek Ministry of National Education and the Ethics Committee of Harokopio University of Athens, and the study was conducted in accordance with the ethical standards specified in the 1964 Declaration of Helsinki. Parents who agreed to the participation of their children in the study had to sign the consent form and provide their contact details. |
| HUNT | The study is approved by the Regional Committee for Medical and Health Research Ethics and by the Norwegian Data Protection Authority, and all study participants gave consent to take part in the study. |
| Piccolipiù | The protocol of the study has been approved by the Ethics committees of the Local Health Unit Roma E (management centre), of the Istituto Superiore di Sanità (National Institute of Public Health) and of each local centre. Standard procedures for the protection of confidential individual information were applied according to the Italian law. Consent forms for participation was signed by the mother and also by the father, when both legally responsible for the newborn. |
| SWS | SWS study was conducted according to the guidelines laid down in the Declaration of Helsinki and was approved by the Southampton and South West Hampshire Local Research Ethics Committee (06/Q1702/104). Written informed consent was obtained from all participating women and by a parent or guardian with parental responsibility on behalf of their children. |

### **Supplementary Text 1.** Description of general characteristics, consent, and ethical approvals in the cohort studies

**Amsterdam Born Children and their Development Study (ABCD)**

Between January 2003 and March 2004, all pregnant women living in Amsterdam were asked to participate in the ABCD study during their first prenatal visit to an obstetric care provider (general practitioner, midwife, or gynaecologist) (1). Of the 12 373 women approached, 8266 women filled out the pregnancy questionnaire (response rate: 67%). Of this group, 7050 women granted permission for follow-up (85%) and 7043 women granted permission for perusal of her and her child’s medical files (85%). Through a questionnaire, women provided information on time to pregnancy (in months) and mode of conception. Blood pressure, heart rate, and blood-based biomarkers (all except insulin and HbA1c) were measured at two timepoints: ages 5-6 years and 11 years. After a 10-minute rest period to relax the child, blood pressure and heart rate were measured twice on the right arm, with the arm supported at heart level and the child in a sitting position, using an Omron 705 IT (Omron Heathcare Inc) with appropriate cuff size (arm circumference, 17–22 cm); mean SBP, DBP, and HR were calculated. Biomarkers were determined based on fasting capillary blood samples that were collected using a validated ambulatory collection kit (Demecal: Lab Anywhere, Haarlem, the Netherlands).

Up to 61 ART-conceived offspring and 4,701 NC offspring were included in this study (singleton births only). ABCD contributed results to the main meta-analysis (ART vs. NC) and to the additional meta-analysis stratified by sex, sub-fertility, and IVF/ICSI, for all study outcomes, except insulin and HbA1c. ABCD also contributed to the multicohort trajectory analysis. Data were available for all study confounders (i.e., maternal age, parity, BMI, smoking, education, ethnicity and offspring sex and age at outcome assessment). Confounders were measured by questionnaire (maternal self-report) administered during first trimester of pregnancy.

Approval for the ABCD study was obtained from the Central Committee on Research involving Human Subjects in the Netherlands, the Medical Ethical Committees of the participating hospitals, and from the Registration Committee of the Municipality of Amsterdam. Written informed consent was obtained from all participating mothers.

**Avon Longitudinal Study of Parents and Children (ALSPAC)**

ALSPAC is a prospective birth cohort study that recruited all pregnant women residing within the catchment area of 3 National Health Service authorities in southwest England with an expected date of delivery between April 1991 and December 1992 (2-4). The initial number of pregnancies enrolled is 14,541 (for these at least one questionnaire has been returned or a “Children in Focus” clinic had been attended by 19/07/99). Of these initial pregnancies, there was a total of 14,676 fetuses, resulting in 14,062 live births and 13,988 children who were alive at 1 year of age. Please note that the study website contains details of all the data that is available through a fully searchable data dictionary and variable search tool" and reference the webpage: <http://www.bristol.ac.uk/alspac/researchers/our-data/>. Detailed information has been collected from offspring and their parents using questionnaires, data extraction from medical records, linkage to health records, and clinic assessments up to the last completed contact.

Up to 12 repeated SBP/DBP/HR measurements, 7 repeated lipids measurements (both from approx. age 3 to 26 years), and 4 repeated glucose measures from age 7 to 26 years were collected from the ALSPAC offspring at dedicated research clinics. At each clinic SBP, DBP and HR were measured at least twice on the right arm after a period of rest, with the child sitting at rest with the arm supported. Blood pressure and heart rate measures were taken using a validated device, and the mean of two or three measures was used. Blood biomarkers were obtained using standard clinical chemistry assays based on non-fasting plasma samples up to age 9, and using fasting samples at older ages, except for non-fasting glucose at age seven which was measured as part of metabolic trait profiling, using Nuclear Magnetic Resonance (NMR) spectroscopy. Lipid quantification (total cholesterol, low-density lipoprotein cholesterol, high-density lipoprotein cholesterol, and triglycerides) was performed according to the standard Lipid Research Clinics Protocol using enzymatic reagents for determining HDL and triglycerides concentrations. LDL levels were calculated by applying the Freidewald equation.

Up to 56 offspring conceived by ART and 10,380 NC offspring were included in this study (including multiple births). ALSPAC contributed results to the main meta-analysis (ART vs. NC) and to the two additional meta-analysis stratified by sex and sub-fertility, for all study outcomes. ALSPAC also contributed to the multicohort trajectory analysis. ALSPAC data were available for all study confounders (i.e., maternal age, parity, BMI, smoking, education, ethnicity and offspring sex and age at outcome assessment).

Ethical approval for the study was obtained from the ALSPAC Ethics and Law Committee and the Local Research Ethics Committees. Informed consent for the use of data collected via questionnaires and clinics was obtained from participants following the recommendations of the ALSPAC Ethics and Law Committee at the time. At age 18, study children were sent 'fair processing' materials describing ALSPAC’s intended use of their health and administrative records and were given clear means to consent or object via a written form. Data were not extracted for participants who objected, or who were not sent fair processing materials. Ethical approval for the study was obtained from the ALSPAC Law and Ethics committee and local research ethics committees (NHS Haydock REC: 10/H1010/70).

**Babies After SCOPE: Evaluating the Longitudinal Impact on Neurological and Nutritional Endpoints (BASELINE)**

The Cork BASELINE Birth Cohort Study (5) is the first Irish prospective birth cohort study and provides detailed information on maternal health, fetal growth, childhood nutrition, growth, and development in the first five years of life. Participants were healthy nulliparous women with singleton pregnancies recruited from the Screening for Pregnancy Endpoints (SCOPE) pregnancy cohort. Detailed information, including information on demographics, lifestyle, obstetric history (including history of fertility and ART) and maternal anthropometric assessment were collected by research midwives at 15 weeks and 20 weeks’ gestation using questionnaires and clinical examination. Child anthropometric measures were collected at each visit according to standard operating procedures. Blood pressure was measured at 2 timepoints, at mean ages 2.2 and 5.1 years, and heart rate was measured at one timepoint, at age 5.1 years. Measurements were taken by trained nurses using the Dinamap V100, in accordance with manufacturer’s instructions. Blood pressure was taken while sitting and relaxed. Three recordings were taken using the Dinamap V100, with an appropriate child-sized cuff. The average systolic was calculated and the average diastolic was calculated and stored on a centralized, secure internet-based database. The child’s heart rate was also recorded on the Dinamap V100 in accordance with manufacturer’s instructions and recorded on the database.

Up to 20 ART-conceived offspring and 1,031 NC offspring were included in this study (singleton births only). BASELINE contributed results to the main meta-analysis (ART vs. NC) and to the additional meta-analysis stratified by sex and sub-fertility, for blood pressure, and heart rate. Data were available for most study confounders (maternal age, BMI, smoking, education, ethnicity and offspring sex and age at outcome assessment), except parity as only nulliparous women were included.

Research objectives and measurements in this birth cohort were conducted according to the guidelines laid down in the Declaration of Helsinki and all procedures were approved by the Clinical Research Ethics Committee of the Cork Teaching Hospitals, [ref ECM5(9) 01/07/ 2008]. Families provided written informed consent at 20 weeks’ gestation or at birth to participate in BASELINE follow-up.

**Barwon Infant Study (BIS)**

BIS is a prospective pre-birth population-based cohort study (n = 1064 mother–1074 infant pairs [10 sets of twins]) with antenatal recruitment conducted in the Barwon region in Victoria, Australia(6). Pregnant women were recruited before 28 weeks of gestation between years 2010 and 2013. Detailed questionnaire and clinical data and extensive biospecimens have been collected from multiple time points from pregnancy to 4 years of age. Blood pressure, and heart rate were measured once at mean age 4.2 years, and blood-based biomarkers (all except insulin and HbA1c) were measured at two timepoints, at mean ages 1 year and 4.2 years.

Up to 35 offspring conceived by ART and 673 NC offspring were included in this study. BIS contributed results to the main meta-analysis (ART vs. NC) and to additional sex-stratified meta-analysis, for all outcomes except insulin and HbA1c. BMI was calculated from height and weight measured by research staff during clinic visits. Body fat percentage was measured using DEXA scanning at 4 years of age. Data were available for all study confounders (i.e., maternal age, parity, BMI, smoking, education, ethnicity and offspring sex and age at outcome assessment).

Ethics approval was obtained from the Barwon Health Human Research Ethics Committee (10/24). All mothers provided written informed consent.

**Clinical review of the Health of 22–33 years old conceived with and without ART (CHART)**

The CHART study (7) is a clinical review of a cohort comprising 547 ART-conceived adults and 549 matched naturally conceived (NC) controls. Recruitment was by letter (postal mailing), with a follow-up letter after 3 weeks and a phone call after a further 3 weeks. Additional attempts at contacting the participants included the use of social media and phone calls to their mothers. Data on clinical and biomarker outcomes were measured including cardiovascular structure and function, auxology, respiratory function, cardiometabolic profile and epigenome-wide DNA methylation analysis. Confounding variables were collected via a questionnaire.

Up to 130 ART-conceived and 73 NC offspring were included in this study. CHART contributed results to the main meta-analysis (ART vs. NC) and to additional meta-analysis stratified by sex and ET/FET, for all study outcomes except HbA1c. Data were available for some confounders (maternal age, parity, education, and offspring sex and age at outcome assessment), but not maternal BMI, smoking or ethnicity.

The study was approved by the Royal Children's Hospital Human Research Ethics Committee, and all study participants provided consent to take part in the study.

**Etude de cohorte généraliste, menée en France sur les Déterminants pré et post natals précoces du développement psychomoteur et de la santé de l’Enfant (EDEN)**

EDEN is a birth cohort study that enrolled 2,002 pregnant women attending their prenatal visit before 24 weeks' gestation at Nancy and Poitiers university hospitals (France) between 2003 and 2006 (8). Detailed information has been collected from parents using questionnaires (including on the mode of conception and fertility treatment), data extraction from obstetrical file and three clinic assessments up to the last completed contact at age 11 yrs. Blood pressure and heart rate were measured with an appropriately sized cuff and an oscillometer COLIN 8800 C device, according to a standardized protocol. Children rested, lying down for 5 minutes, and remained in that position and their blood pressure was measured on the left arm three times at 2-minute intervals, with the mean of the three measurements used. Plasma/serum samples were taken from children at mean 5.7 years. Samples were stored in freezers with alarm control at -80 degree Celsius.

Up to 22 ART-conceived offspring and 1,362 NC offspring were included in this study. EDEN contributed results to the main meta-analysis (ART vs. NC) and to additional meta-analysis stratified by sex and sub-fertility, for all study outcomes except HbA1c. Data were available for all study confounders (maternal age, BMI, smoking, education, ethnicity, parity and offspring sex and age at outcome assessment).

The study received approval from the ethics committee (CCPPRB) of Kremlin Bicêtre on 12 December 2002 and from CNIL (Commission Nationale Informatique et Liberté), the French data privacy institution. All subjects gave their informed consent for inclusion before they participated in the study. Consent for the child was obtained from both parents after the child's birth.

**Gene and Environment: Prospective Study on Infancy in Italy (GASPII)**

GASPII is a prospective birth cohort study of 700 children born in selected maternal units located in Rome (9).

Up to 8 ART-conceived offspring and 554 NC offspring were included in this study. GASPII contributed results to the main meta-analysis (ART vs. NC) to additional meta-analysis stratified by sub-fertility, for all outcomes, except HbA1c. Data were available for all study confounders (maternal age, BMI, smoking, education, ethnicity, parity and offspring sex and age at outcome assessment).

The protocol of the study has been approved by the Ethics committees of the Università Cattolica del Sacro Cuore, Rome, and all study participants provided consent to take part in the study.

**Generation R (Gen R)**

Gen R is a population-based prospective cohort study from fetal life until adulthood(10, 11). In total, 9,778 mothers with a delivery date from April 2002 until January 2006 were enrolled in the study. Response at baseline was 61%. Extensive assessments are performed in mothers, fathers, and their children. Blood pressure, heart rate, and all biomarkers (except HbA1c) were measured at 2 timepoints at mean ages 6.2 years and 9.8 years. Blood pressure and heart rate were measured at the right brachial artery, 4 times with 1-minute intervals, using a validated automatic sphygmomanometer Datascope Accutor Plus TM (Paramus, NJ). The mean values of the last 3 measurements were used. 30-minute fasting venous blood samples were obtained and used to measure biomarkers. Lipids levels were measured with enzymatic methods using the Cobas 8000 analyzer (Roche, Almere, The Netherlands). Quality control demonstrated intra- and inter-assay coefficients of variation ranging from 0.77 to 1.39%, and from 0.87 to 2.40%, respectively.

Up to 47 ART-conceived offspring and 4,334 NC offspring were included in this study. Gen R contributed results to the main meta-analysis (ART vs. NC) to additional meta-analysis stratified by sex, for all outcomes except HbA1c. Data were available for all study confounders (maternal age, BMI, smoking, education, ethnicity, parity and offspring sex and age at outcome assessment).

The study has been approved by the Medical Ethical Committee of the Erasmus MC, University Medical Center in Rotterdam (MEC-2012-165-NL40020.078.12). Written informed consent was obtained from the parents or legal representatives of the children. Even with consent of the parents, when the child is not willing to participate actively, no measurements are performed.

**Generation XXI (G21)**

Generation XXI (G21) is a prospective population-based birth cohort that recruited pregnant women delivering live-born infants (including multiple births) between April 2005 and August 2006 at all five public maternity units that served the metropolitan area of Porto, Portugal(12). Overall, 8,647 infants with gestational age above 23 weeks and their mothers (n=8,495) were enrolled (91.4% participation). Subsequent evaluations of the entire cohort took place when children were four (n=7,459), seven (n=6,889) and ten (n=6,397) years old and 13 (n=4,640, interrupted due to the COVID-19 pandemic). The cohort has more than 95% Caucasian participants. Data on demographic and socioeconomic characteristics, lifestyles, obstetric history, and anthropometrics were collected within 72 hours after delivery, in a face-to-face interview conducted by trained interviewers using structured questionnaires. During follow-up, trained researchers performed anthropometric and blood pressure measurements and obtained a fasting blood sample, according to standard procedures (13). All outcomes were measured at least during over follow-up, blood pressure was measured at three timepoints. Two measurements of systolic blood pressure (SBP) and diastolic blood pressure (DBP), separated by at least 5 minutes, were taken after a 10-minute rest. If the difference between them was lower than 5 mm Hg for both SBP and DBP, the mean was calculated; if the difference was larger than 5 mm Hg, a third measurement was taken, and the mean of the two closest values was used. After an overnight fast, a venous blood sample was collected before 11 a.m. Samples were centrifuged at 3,500 rpm for 10 minutes and then stored at −80°C. Glucose was measured using an ultraviolet enzymatic assay (hexokinase method), insulin using an electrochemiluminescence immunoassay, triglycerides, and total and high-density lipoprotein cholesterol (HDL cholesterol) using an enzymatic colorimetric assay, LDL cholesterol was calculated using the Friedewald equation.

Up to 92 ART-conceived offspring and 5746 NC offspring were included in this study (including multiple births). G21 contributed results to the main meta-analysis (ART vs. NC) to additional meta-analysis stratified by sex, sub-fertility, and IVF/ICSI, for all study outcomes. G21 also contributed to the multicohort trajectory analysis. Data were available for all study confounders (maternal age, BMI, smoking, education, maternal country of birth, parity and offspring sex and age at outcome assessment).

The Ethics Committee of Hospital de São João, and of the Institute of Public Health of the University of Porto approved the study protocols. The study complies with the Ethical Principles expressed in the Helsinki Declaration and with the national legislation and was registered with the Portuguese Authority for Data Protection. In all evaluations, participants were informed about the purposes and design of the study, as well as the potential discomfort caused by participation. Signed informed consent was obtained from all parents or legal guardians, and oral assent was obtained from children at each evaluation.

**Growing up in Singapore Towards healthy Outcomes (GUSTO)**

GUSTO recruited pregnant women aged 18 years and above, attending their first trimester antenatal dating ultrasound scan clinic at Singapore’s two major public maternity units(14). Women were eligible if 18 years and older, Singaporean citizens or permanent residents, with self-reported homogenous ethnic ancestry (Chinese, Indian, Malay), intended to deliver at the either of the recruitment hospitals and reside in Singapore for the next 5 years. Women greater than 14 weeks of gestation, receiving chemotherapy, psychotropic medications, or having an existing type I diabetes mellitus diagnosis at the time of recruitment were excluded. Women who ultimately did not agree to donate birth tissues (cord, placenta, cord blood) were also excluded. Women were asked to self-report whether the current pregnancy was conceived via IVF and use of assisted reproductive technologies, along with relevant treatment modalities, were confirmed via medical record review by a senior obstetrician and fertility consultant. Women reporting IVF conception with multiple gestations were further excluded.

Maternal obstetric and medical history including self-reported pre-pregnancy body weight, sociodemographic characteristics, and health behaviors, such as personal and family tobacco smoking, were ascertained by study staff administered standardized questionnaire at recruitment and at a study visit at 26-28 weeks gestation. Mode of delivery, procedures, and complications and birth weight, length, and head circumference were abstracted from delivery record. Child blood pressures were measured at 3-, 4-, 5-, and 6-year visits using a DINAMAP CARESCAPE V100 (GE Healthcare, Milwaukee, USA) automated blood pressure monitor. Children were fitted with an appropriately sized cuff (8–13 cm or 12–19 cm; GE CRITIKON) on the bare, upper arm and, after a 5-min initial rest in the seated position, measured twice with 25–30 s between measurements. If either systolic or diastolic blood pressure varied more than 10 mmHg, a third measure was taken. A simple mean was taken of all valid, repeated measures for a subject (15). At age 6, children were asked to fast the evening before the study visit. At the visit, venepuncture was performed by study staff and a peripheral blood sample was spun, aliquoted, and stored at −80 °C. One plasma aliquot was immediately assayed for glucose concentrations as reported above for maternal glucose. In 2019, one serum aliquot per child was thawed and analyzed for the following biomarkers at the College of American Pathologist-accredited NUH Referral Laboratory (Singapore) following standard clinical laboratory protocols:

Up to 66 ART-conceived offspring and 935 NC offspring were included in this study (singletons only). GUSTO contributed results to the main meta-analysis (ART vs. NC) to additional meta-analysis stratified by sex, IVF/ICSI, and ET/FET for all study outcomes except HbA1c. Data were available for all study confounders (maternal age, BMI, smoking, education, ethnicity, parity and offspring sex and age at outcome assessment).

Study protocols following the principles of the Declaration of Helsinki and were approved by the respective ethics committees for two hospitals: National Healthcare Group Domain Specific Review Board (NUH) and SingHealth Centralized Institutional Review Board (KKH). All participants in this study provided informed consent to participate and contribute their data to publications. The GUSTO study is registered under study ID: NCT01174875 (clinicaltrials.gov) which broadly covers investigations of parental and gestational influences on child health.

**Healthy Growth Study (HGS)**

HGS is a child cohort study started in 2007 that recruited schoolchildren aged 9–13 years, attending primary schools located in municipalities within the counties of Attica, Aitoloakarnania, Thessaloniki and Iraklio, in Greece(16). Participants underwent a physical examination by two trained members of the research team. The protocol and equipment used were the same in all schools. Data on the socio-economic background of the families having at least one child participating in the study were collected from the parents (most preferably from the mother) during scheduled face-to-face interviews at school. All subjects underwent BP assessment using a mercury sphygmomanometer, which was placed in the right arm with the subject seated and quiet after a 5-min rest. Assessment of the mid-upper arm circumference was performed before the blood pressure screening and different cuff sizes were used as appropriate. The 1st and 5th Korotkoff sound were used for the identification of the systolic and diastolic BP level. Two consecutive measurements were performed for each child, with a 2-min interval and if the pressure readings differed by more than 10 mmHg, additional measurement was carried out. Mean value from two or three consecutive readings of SBP and DBP taken from each child was used. A member of the research team contacted all parents and children the day before the blood tests in order to make sure that they would follow the overnight fast. Subjects visited laboratory in the morning and submitted to hematologic screening test. Professional staff collected blood samples (maximum 23 ml of blood for each participant) conducting venipuncture (17)

Up to 63 ART-conceived offspring and 2,182 NC offspring were included in this study. HGS contributed results to the main meta-analysis (ART vs. NC) to additional meta-analysis stratified by sex, and IVF/ICSI, for all study outcomes except HbA1c. Data were available for all study confounders (maternal age, BMI, smoking, education, ethnicity, parity and offspring sex and age at outcome assessment).

Approval to conduct the study was granted by the Greek Ministry of National Education and the Ethics Committee of Harokopio University of Athens, and the study was conducted in accordance with the ethical standards specified in the 1964 Declaration of Helsinki. Parents who agreed to the participation of their children in the study had to sign the consent form and provide their contact details.

**Piccolipiù**

Piccolipiù is a prospective birth cohort study of 3358 children born in selected maternal units located in five Italian cities (Florence, Rome, Trieste, Turin, and Viareggio)(18) between 2011-2015. Piccolipiù study recruited singleton pregnant women aged at least 18 years old and giving birth in one of the selected maternity units. Mothers were recontacted at 6, 12, 24 and 48 months after delivery for follow-up questionnaires.At 4 years, a clinical examination including blood pressure measurements was carried out.

Up to 86 ART-conceived offspring and 2,479 NC offspring were included in this study. Piccolipiù contributed results to the main meta-analysis (ART vs. NC) to additional meta-analysis stratified by sex, and sub-fertility, for blood pressure outcomes. Data were available for all study confounders (maternal age, BMI, smoking, education, ethnicity, parity and offspring sex and age at outcome assessment).

The protocol of the study has been approved by the Ethics committees of the Local Health Unit Roma E (management centre), of the Istituto Superiore di Sanità (National Institute of Health) and of each local centre. Standard procedures for the protection of confidential individual information were applied according to the Italian law. Consent forms for participation was signed by the mother and also by the father, when both legally responsible for the newborn.

**Southampton Women's Survey (SWS)**

SWS is a population-based prospective birth cohort study of 12 583, initially non-pregnant, women aged 20–34 years, living in the city of Southampton, UK(19). Assessments of lifestyle, diet and anthropometry were done at study entry in 1998–2002. Women who subsequently became pregnant with singleton pregnancies were followed up during pregnancy; and their offspring have been studied in infancy and childhood. Information on ART was obtained at the time of the first scan by questioning the mother. SBP, DBP and HR of the children were measured using a Dinamap

Critikon 8100 monitor.

Up to 36 ART-conceived offspring and 2,554 NC offspring were included in this study (singleton births only). SWS contributed results to the main meta-analysis (ART vs. NC) to additional meta-analysis stratified by sex, for blood pressure and heart rate outcomes. Data were available for all study confounders (maternal age, BMI, smoking, education, ethnicity, parity and offspring sex and age at outcome assessment).

The SWS was conducted according to the guidelines laid down in the Declaration of Helsinki and was approved by the Southampton and South West Hampshire Local Research Ethics Committee (06/Q1702/104). Written informed consent was obtained from all participating women and by a parent or guardian with parental responsibility on behalf of their children.

**The Trøndelag Health Study (HUNT)**

The Trøndelag Health Study (HUNT) is a population-based study where all adult residents of the Nord-Trøndelag region, Norway have been invited to repeated surveys since the 1980s. Since the 1990s, all adolescents (aged 13-19 years) in the region have also been invited (the Young-HUNT Study)(20, 21). The participants have consented to data linkage to health registries, such as the Medical Birth Registry of Norway (MBRN), which includes information on virtually all births in Norway since 1967. In this study, we included participants from the Young-HUNT1 (1995-97), Young-HUNT2 (1999-2000) and Young-HUNT3 (2006-08) surveys, which included measurements of SBP and DBP. Information on mode of conception was obtained through linkage to information from the MBRN.

Up to 121 ART-conceived offspring and 9,711 NC offspring were included in this study (including multiple births). HUNT contributed results to the main meta-analysis (ART vs. NC) to additional meta-analysis stratified by sex, IVF/ICSI, and ET/FET, for blood pressure outcomes. Data were available for some study confounders (maternal age, parity and offspring sex and age at outcome assessment), but not maternal BMI, smoking, education, or ethnicity, but ~97% of the population in this region was of European ancestry at the time of HUNT2 (21, 22).

The study is approved by the Regional Committee for Medical and Health Research Ethics and by the Norwegian Data Protection Authority.

### **Supplementary Text 2**. Cohort specific acknowledgments/funding and data access

**Avon Longitudinal Study of Parents and Children (ALSPAC)**

We are extremely grateful to all of the families who took part in ALSPAC, the midwives for their help in recruiting them, and the whole ALSPAC team, which includes interviewers, computer and laboratory technicians, clerical workers, research scientists, volunteers, managers, receptionists and nurses.

Core funding for the Avon Longitudinal Study of Parents and Children (ALSPAC) is provided by the UK Medical Research Council and Wellcome (217065/Z/19/Z) and the University of Bristol. A comprehensive list of grants funding is available on the ALSPAC website (http://www.bristol.ac.uk/alspac/external/documents/grant-acknowledgements.pdf). DAL and AE work in a unit that is supported by the University of Bristol and UK Medical Research Council (MC_UU_00011/6) and DAL holds a European Research Council Advanced Grant (ERC grant agreement no 669545) and is a NIHR Senior Investigator (NF-0616-10102). The funders had no role in the design of the study, the collection, analysis, or interpretation of the data; the writing of the manuscript, or the decision to submit the manuscript for publication. The views expressed in this paper are those of the authors and not necessarily those of any funder.

Researchers interested in accessing the ALSPAC data used in this study will need to submit a research proposal ( https://proposals.epi.bristol.ac.uk/) for consideration by the ALSPAC Executive Committee.

**Amsterdam Born Children and their Development Study (ABCD)**

We are grateful to all participating hospitals, obstetric clinics, and general practitioners for their assistance in implementing the ABCD study and thank all of the women who participated for their cooperation. Core funding of the ABCD-study is provided by the Academic Medical Centre, Amsterdam, the Public Health Services, Amsterdam, and the Dutch Organization for Health Research and Development (ZonMw).

Researchers interested in accessing the ABCD data used in this study will need to submit a short research proposal that should include information on the background, research questions and methods, a plan for publication, timetable and budget. For more information, please see the ABCD website or contact one of the principal investigators at.

**Babies After SCOPE: Evaluating the Longitudinal Impact on Neurological and Nutritional Endpoints (BASELINE)**

The authors thank the families for their continued support and the Cork BASELINE Birth Cohort Study research team. SCOPE Ireland was supported by the Health Research Board, Ireland (CSA 2007/2). The BASELINE cohort was funded by the National Children’s Research Centre, Dublin, Ireland, and the Food Standards Agency of the United Kingdom (grant no. TO7060).

Researchers interested in accessing the BASELINE data used in this study can contact []. Further information be found at [http://www.baselinestudy. net/ or <http://www.birthcohorts.net/>].

**Barwon Infant Study (BIS)**

We thank the BIS participants for the generous contribution they have made to this project. We also thank current and past staff for their efforts in recruiting and maintaining the cohort and in obtaining and processing the data and biospecimens.

The establishment work and infrastructure for the BIS was provided by the Murdoch Children’s Research Institute, Deakin University and Barwon Health. Subsequent funding was secured from the National Health and Medical Research Council of Australia, The Jack Brockhoff Foundation, the Scobie Trust, the Shane O’Brien Memorial Asthma Foundation, the Our Women’s Our Children’s Fund Raising Committee Barwon Health, The Shepherd Foundation, the Rotary Club of Geelong, the Ilhan Food Allergy Foundation, GMHBA Limited and the Percy Baxter Charitable Trust, Perpetual Trustees. In-kind support was provided by the Cotton On Foundation and CreativeForce. Research at Murdoch Children’s Research Institute is supported by the Victorian Government's Operational Infrastructure Support Program. This work was also supported by NHMRC Senior Research Fellowships (1064629 to DB; 1045161 to RS) and NHMRC Investigator Grants to DB (1175744).

Researchers interested in accessing the BIS data used in this study will need to make a request for consideration by the BIS Steering Committee. Further information about BIS can be obtained via

**Clinical review of the Health of 22–33 years old conceived with and without ART (CHART)**

We would like to acknowledge the participants who generously gave their time to the study and the invaluable contribution of Ms. Jane Koleff to the development of the protocol and in the training of all assessors to undertake the clinical assessments.

The CHART study was supported by the Victorian State Government Operational Infrastructure Support and the Australian Government NHMRC IRIISS awarded to the Murdoch Children’s Research Institute, and funded by a National Health & Medical Research Council Project Grant (APP1099641; 2016–2017), Royal Children’s Hospital Research Foundation, Monash IVF Research and Education Foundation, and Reproductive Biology Unit Sperm Fund, Melbourne IVF

Researchers interested in accessing the CHART data used in this study should.

**Etude de cohorte généraliste, menée en France sur les Déterminants pré et post natals précoces du développement psychomoteur et de la santé de l’Enfant (EDEN)**

The authors thank the cohort participants and the EDEN mother-child study group, whose members are: I. Annesi-Maesano, J.Y. Bernard, J. Botton, M.A. Charles, P. Dargent-Molina, B. de Lauzon-Guillain, P. Ducimetière, M. de Agostini, B. Foliguet, A. Forhan, X. Fritel, A. Germa, V. Goua, R. Hankard, B. Heude, M. Kaminski, B. Larroque†, N. Lelong, J. Lepeule, G. Magnin, L. Marchand, C. Nabet, F Pierre, R. Slama, M.J. Saurel-Cubizolles, M. Schweitzer, O. Thiebaugeorges.

The EDEN study was supported by Foundation for medical research (FRM), National Agency for Research (ANR), National Institute for Research in Public health (IRESP: TGIR cohorte santé 2008 program), French Ministry of Health (DGS), French Ministry of Research, INSERM Bone and Joint Diseases National Research (PRO-A) and Human Nutrition National Research Programs, Paris-Sud University, Nestlé, French National Institute for Population Health Surveillance (InVS), French National Institute for Health Education (INPES), the European Union FP7 programmes (FP7/2007- 2013, HELIX, ESCAPE, ENRIECO, Medall projects), Diabetes National Research Program (through a collaboration with the French Association of Diabetic Patients (AFD)), French Agency for Environmental Health Safety (now ANSES), Mutuelle Générale de l’Education Nationale a complementary health insurance (MGEN), French national agency for food security, French speaking association for the study of diabetes and metabolism (ALFEDIAM).

Researchers interested in accessing the EDEN data used in this study EDEN data should email

] and [] and complete a dedicated project form for evaluation by the EDEN steering committee. Further details on the study can be found on the EDEN website: [https://eden.vjf.inserm.fr/ index.php?lang¼en]

**Generation R (Gen R)**

The authors gratefully acknowledge the contribution of participants, research collaborators, general practitioners, hospitals, midwives, and pharmacies in Rotterdam.

The general design of the Generation R Study is made possible by financial support from the Erasmus MC, University Medical Center, Rotterdam, Erasmus University Rotterdam, Netherlands Organization for Health Research and Development (ZonMw), Netherlands Organisation for Scientific Research (NWO), Ministry of Health, Welfare and Sport and Ministry of Youth and Families. This project received funding from the European Union's Horizon 2020 research and innovation programme (LIFECYCLE, grant agreement No 733206, 2016, European Joint Programming Initiative “A Healthy Diet for a Healthy Life” (JPI HDHL, EndObesity project, ZonMW the Netherlands no. 529051026). RG received funding of the Dutch Heart Foundation (grant number 2017T013), the Dutch Diabetes Foundation (grant number 2017.81.002), and the Netherlands Organization for Health Research and Development (NWO, ZonMW, grant number 543003109). The study sponsors had no role in the study design, data analysis, interpretation of data, or writing of this report.

Researchers interested in accessing the Gen R data used in this study should contact Vincent Jaddoe. Requests will be discussed in the Generation R Study Management Team regarding their study aims, overlap with ongoing studies, logistic consequences and related finances.

**Generation XXI (G2I)**

G21 was funded by Programa Operacional de Saúde – Saúde XXI, Quadro Comunitário de Apoio III and Administração Regional de Saúde Norte (Regional Department of Ministry of Health) and by Foundation for Science and Technology – FCT (UIDB/04750/2020 - Unidade de Investigação em Epidemiologia (EPIUnit), Instituto de Saúde Pública da Universidade do Porto). The funders had no role in study design, data collection and analysis, interpretation of data, or writing of this report.

Researchers interested in accessing the G21 data used in this study should contact Henrique Barros.

**Growing up in Singapore Towards healthy Outcomes (GUSTO)**

We thank the GUSTO study group and all clinical and home-visit staff involved. The voluntary participation of all participants is greatly appreciated.

The GUSTO study group includes. Allan Sheppard,Amutha Chinnadurai, Anne Ferguson-Smith, Anne Eng Neo Goh, Arijit Biswas, Audrey Chia, Birit Leutscher-Broekman, Borys Shuter, Shirong Cai, Cheryl Ngo, Chai Kiat Chng, Shang Chee Chong, Christiani Jeyakumar Henry, Mei Chien Chua, Cornelia Yin Ing Chee, Yam Thiam Daniel Goh, Dennis Bier, Chun Ming Ding, Doris Fok, Eric Andrew Finkelstein, Fabian Kok Peng Yap, George Seow Heong Yeo, Wee Meng Han, Helen Chen, Hugo P S Van Bever, Hazel Inskip, Iliana Magiati, Inez Bik Yun Wong, Jeevesh Kapur, Jenny L Richmond, Jerry Kok Yen Chan, Joshua J Gooley, Krishnamoorthy Niduvaje, Bee Wah Lee, Yung Seng Lee, Leher Singh, Sok Bee Lim, Lourdes Mary Daniel, Seong Feei Loh, Yen-Ling Low, Pei-Chi Lynette Shek, Marielle Fortier, Mark Hanson, Mary Foong-Fong Chong, Michael Meaney, Susan Morton, Wei Wei Pang, Pratibha Agarwal, Anqi Qiu, Boon Long Quah, Rob M van Dam, David Stringer, Salome Antonette Rebello, Wing Chee So, Chin-Ying Hsu, Lin Lin Su, Jenny Tang, Kok Hian Tan, Soek Hui Tan, Oon Hoe Teoh, Victor Samuel Rajadurai, PC Wong and Sudhakar K Venkatesh

Researchers interested in accessing the GUSTO data used in this study should contact Jonathan Huang.

**Healthy Growth Study (HGS)**

The HGS was co-funded by the European Union (European Social Fund – ESF) and Greek national funds through the Operational Program "Education and Lifelong Learning" of the National Strategic Reference Framework (NSRF) - Research Funding Program: Heracleitus II. Investing in knowledge society through the European Social Fund.

Researchers interested in accessing the HGS data used in this study should contact George Moschonis.

**The Trøndelag Health Study (HUNT)**

The Trøndelag Health Study (HUNT) is a collaboration between HUNT Research Centre (Faculty of Medicine and Health Sciences, NTNU, Norwegian University of Science and Technology), Trøndelag County Council, Central Norway Regional Health Authority, and the Norwegian Institute of Public Health. The study was partly funded by the Norwegian Research Council’s Centres of Excellence funding scheme, project no. 262700.

**Piccolipiù**

The authors thank all the families who took part in the study, and the Piccolipiù research group.

The study was funded by the Italian National Centre for Disease Prevention and Control (CCM grant 2010), by the Italian Ministry of Health (art 12 and 12bis Dl .gs. vo 502/92).

**Prospective Study on Infancy in Italy (GASPII)**

The authors are grateful to all the participating families in Rome who take part in this on-going cohort study, and to all the field workers and interviewers of the project. The study was funded by Italian Ministry of Health.

**Southampton Women's Survey (SWS)**

The authors are grateful to the women and children from Southampton who gave their time to take part in the Southampton Women’s Survey and to the research nurses and other staff who collected and processed the data. The SWS is supported by grants from the Medical Research Council, National Institute for Health Research Southampton Biomedical Research Centre, British Heart Foundation, UK Food Standards Agency, British Lung Foundation, Versus Arthritis, University of Southampton and University Hospital Southampton National Health Service Foundation Trust, and the European Union’s Seventh Framework Programme (FP7/2007-2013), project Early Nutrition (grant 289346) and from the European Union's Horizon 2020 research and innovation programme (LIFECYCLE, grant agreement No 733206). Study participants were drawn from a cohort study funded by the Medical Research Council and the Dunhill Medical Trust.

Researchers interested in accessing the SWS data used in this study should contact should apply to Professor Janis Baird with details about the proposed use. Approval would then be required from the SWS Oversight Group.

### **Supplementary Text 3.** Supplementary references

### **Supplementary Table 1**. Descriptive characteristics of the included cohorts in terms of the outcomes, age at outcome assessment, and numbers included in the main analysis and each additional analysis.

| **Cohort / outcome** | **Mean (SD) age** | **Mean (SD) outcome** | **N-ART** | **N-NC** | **N-NC: subfertile** | **N-NC: fertile** | **N-ART: males** | **N-NC: males** | **N-ART: females** | **N-NC: females** | **N-IVF** | **N-ICSI** | **N-fresh ET** | **N-FET** |
| --- | --- | --- | --- | --- | --- | --- | --- | --- | --- | --- | --- | --- | --- | --- |
| **1. ABCD** |  |  |  |  |  |  |  |  |  |  |  |  |  |  |
| 1. SBP | 5.7 (0.5) | 97.7 (8.7) | 25 | 2835 | 742 | 2093 | 12 | 1419 | 12 | 1416 | 8 | 17 | - | - |
| 2. DBP | 5.7 (0.5) | 58.2 (8.1) | 24 | 2835 | 742 | 2093 | 12 | 1418 | 12 | 1416 | 8 | 16 | - | - |
| 3. HR | 5.7 (0.5) | 87.0 (11.1) | 25 | 2860 | 745 | 2115 | 13 | 1433 | 12 | 1427 | 8 | 17 | - | - |
| 4. TC | 5.7 (0.5) | 4.0 (0.7) | 18 | 1894 | 464 | 1430 | 9 | 963 | 9 | 931 | 4 | 14 | - | - |
| 5. HDL-C | 5.7 (0.5) | 1.3 (0.3) | 18 | 1894 | 464 | 1430 | 9 | 963 | 9 | 931 | 4 | 14 | - | - |
| 6. LDL-C | 5.7 (0.5) | 2.3 (0.6) | 18 | 1894 | 464 | 1430 | 9 | 963 | 9 | 931 | 4 | 14 | - | - |
| 7. TG | 5.7 (0.5) | 0.6 (0.3) | 18 | 1894 | 464 | 1430 | 9 | 887 | 9 | 931 | 4 | 14 | - | - |
| 8. Glucose | 5.7 (0.5) | 4.6 (0.5) | 18 | 1894 | 464 | 1430 | 9 | 963 | 9 | 931 | 4 | 14 | - | - |
| **2. ALSPAC** |  |  |  |  |  |  |  |  |  |  |  |  |  |  |
| 1. SBP | 9.9 (0.3) | 103 (9) | 47 | 5732 | - | - | 30 | 2824 | 17 | 2908 | - | - | - | - |
| 10. HbA1c | 9.9 (0.3) | 4.9 (0.3) | 14 | 1292 | - | - | 11 | 657 | 3 | 635 | - | - | - | - |
| 2. DBP | 9.9 (0.3) | 57 (6) | 47 | 5732 | - | - | 30 | 2824 | 17 | 2908 | - | - | - | - |
| 3. HR | 9.9 (0.3) | 79 (11) | 47 | 5733 | - | - | 30 | 2824 | 17 | 2909 | - | - | - | - |
| 4. TC | 9.9 (0.3) | 4.3 (0.7) | 33 | 3872 | - | - | 20 | 1966 | 13 | 1906 | - | - | - | - |
| 5. HDL-C | 9.9 (0.3) | 1.4 (0.3) | 33 | 3872 | - | - | 20 | 1966 | 13 | 1906 | - | - | - | - |
| 6. LDL-C | 9.9 (0.3) | 2.3 (0.6) | 33 | 3871 | - | - | 20 | 1966 | 13 | 1905 | - | - | - | - |
| 7. TG | 9.9 (0.3) | 1.1 (0.6) | 33 | 3872 | - | - | 20 | 1966 | 13 | 1906 | - | - | - | - |
| 8. Glucose | 7.5 (0.3) | 4.2 (0.5) | 27 | 4150 | - | - | 20 | 2145 | 7 | 2005 | - | - | - | - |
| 9. Insulin | 9.9 (0.3) | 13.3 (7.2) | 34 | 3912 | - | - | 20 | 1943 | 13 | 1903 | - | - | - | - |
| **3. BASELINE** |  |  |  |  |  |  |  |  |  |  |  |  |  |  |
| 1. SBP | 2.2 (0.1) | 99.6 (10.1) | 14 | 648 | 95 | 553 | 8 | 330 | 6 | 318 | - | - | - | - |
| 2. DBP | 2.2 (0.1) | 58.2 (6.9) | 14 | 648 | 95 | 553 | 8 | 330 | 3 | 318 | - | - | - | - |
| 3. HR | 5.1 (0.2) | 98.7 (10.6) | 13 | 800 | 96 | 704 | 7 | 400 | 6 | 400 | - | - | - | - |
| **4. BIS** |  |  |  |  |  |  |  |  |  |  |  |  |  |  |
| 1. SBP | 4.2 (0.3) | 106.8 (8.4) | 21 | 506 | - | - | 9 | 266 | 12 | 240 | - | - | - | - |
| 2. DBP | 4.2 (0.3) | 64.2 (6.6) | 21 | 506 | - | - | 9 | 266 | 12 | 240 | - | - | - | - |
| 3. HR | 4.2 (0.3) | 89.7 (9.6) | 21 | 503 | - | - | 9 | 265 | 12 | 238 | - | - | - | - |
| 4. TC | 1.1 (0.07) | 3.3 (0.7) | 30 | 562 | - | - | 14 | 293 | 16 | 269 | - | - | - | - |
| 5. HDL-C | 1.1 (0.07) | 1.2 (0.2) | 30 | 562 | - | - | 14 | 293 | 16 | 269 | - | - | - | - |
| 6. LDL-C | 1.1 (0.07) | 1.1 (0.4) | 30 | 562 | - | - | 14 | 293 | 16 | 269 | - | - | - | - |
| 7. TG | 1.1 (0.07) | 2.8 (1.0) | 30 | 562 | - | - | 14 | 293 | 16 | 269 | - | - | - | - |
| 8. Glucose | 1.1 (0.07) | 2.8 (1.0) | 30 | 550 | - | - | 14 | 290 | 16 | 260 | - | - | - | - |
| **5. CHART** |  |  |  |  |  |  |  |  |  |  |  |  |  |  |
| 1. SBP | 27.4 (2.7) | 122.1 (11.0) | 130 | 73 | - | - | 54 | 25 | 76 | 48 | - | - | 93 | 37 |
| 2. DBP | 27.4 (2.7) | 72.7 (8.0) | 130 | 73 | - | - | 54 | 25 | 76 | 48 | - | - | 93 | 37 |
| 3. HR | 27.4 (2.7) | 63.6 (9.7) | 128 | 71 | - | - | 54 | 24 | 74 | 47 | - | - | 92 | 36 |
| 4. TC | 27.4 (2.7) | 4.7 (0.9) | 128 | 71 | - | - | 54 | 25 | 74 | 46 | - | - | 90 | 38 |
| 5. HDL-C | 27.4 (2.7) | 1.7 (0.5) | 128 | 71 | - | - | 54 | 25 | 74 | 46 | - | - | 90 | 38 |
| 6. LDL-C | 27.4 (2.7) | 2.6 (0.7) | 128 | 71 | - | - | 54 | 25 | 74 | 46 | - | - | 90 | 38 |
| 7. TG | 27.4 (2.7) | 1.0 (0.5) | 128 | 71 | - | - | 54 | 25 | 74 | 46 | - | - | 90 | 38 |
| 8. Glucose | 27.4 (2.7) | 4.7 (0.4) | 124 | 69 | - | - | 52 | 25 | 72 | 44 | - | - | 88 | 36 |
| 9. Insulin | 27.4 (2.7) | 8.5 (5.6) | 126 | 70 | - | - | 53 | 24 | 73 | 46 | - | - | 88 | 38 |
| **6. EDEN** |  |  |  |  |  |  |  |  |  |  |  |  |  |  |
| 1. SBP | 3.2 (0.07) | 92.5 (8.3) | 18 | 1077 | 140 | 825 | 8 | 570 | 11 | 513 | - | - | - | - |
| 2. DBP | 3.2 (0.07) | 49.3 (7.7) | 18 | 1077 | 140 | 825 | 8 | 570 | 11 | 513 | - | - | - | - |
| 3. HR | 3.2 (0.07) | 109.4 (10.5) | 18 | 1071 | 138 | 821 | 8 | 566 | 11 | 511 | - | - | - | - |
| 4. TC | 5.7 (0.1) | 4.3 (0.7) | 8 | 594 | 77 | 454 | 5 | 326 | 3 | 268 | - | - | - | - |
| 5. HDL-C | 5.7 (0.1) | 1.5 (0.3) | 8 | 593 | 77 | 453 | 5 | 326 | 3 | 267 | - | - | - | - |
| 6. LDL-C | 5.7 (0.1) | 2.6 (0.6) | 8 | 592 | 77 | 452 | 5 | 325 | 3 | 267 | - | - | - | - |
| 8. Glucose | 5.7 (0.1) | 4.5 (0.4) | 7 | 524 | 70 | 403 | 5 | 291 | 2 | 233 | - | - | - | - |
| 9. Insulin | 5.7 (0.1) | 14.2 (15.7) | 11 | 676 | 92 | 521 | 6 | 376 | 5 | 300 | - | - | - | - |
| **7. GASPII** |  |  |  |  |  |  |  |  |  |  |  |  |  |  |
| 1. SBP | 7.8 (0.2) | 106.3 (11.7) | 8 | 441 | 40 | 390 | - | - | - | - | - | - | - | - |
| 2. DBP | 7.8 (0.2) | 66.3 (10.5) | 8 | 441 | 40 | 390 | - | - | - | - | - | - | - | - |
| 3. HR | 1.4 (0.1) | 101.1 (19.3) | 7 | 502 | 45 | 448 | - | - | - | - | - | - | - | - |
| 4. TC | 8.7 (0.3) | 4.2 (0.7) | 7 | 381 | 36 | 336 | - | - | - | - | - | - | - | - |
| 5. HDL-C | 8.7 (0.3) | 1.5 (0.4) | 7 | 381 | 36 | 336 | - | - | - | - | - | - | - | - |
| **8. Gen R** |  |  |  |  |  |  |  |  |  |  |  |  |  |  |
| 1. SBP | 6.2 (0.5) | 102.6 (8.2) | 42 | 3969 | - | - | 19 | 1982 | 23 | 1987 | - | - | - | - |
| 2. DBP | 6.2 (0.5) | 60.6 (6.8) | 42 | 3969 | - | - | 19 | 1982 | 23 | 1987 | - | - | - | - |
| 3. HR | 6.2 (0.5) | 83.6 (10.0) | 44 | 4165 | - | - | 19 | 2090 | 25 | 2075 | - | - | - | - |
| 4. TC | 6.2 (0.5) | 4.2 (0.6) | 26 | 2878 | - | - | 9 | 1474 | 17 | 1404 | - | - | - | - |
| 5. HDL-C | 6.2 (0.5) | 1.3 (0.3) | 26 | 2882 | - | - | 9 | 1476 | 17 | 1406 | - | - | - | - |
| 6. LDL-C | 6.2 (0.5) | 2.4 (0.6) | 25 | 2881 | - | - | 9 | 1476 | 17 | 1405 | - | - | - | - |
| 7. TG | 6.2 (0.5) | 1.1 (0.5) | 26 | 2871 | - | - | 9 | 1474 | 17 | 1397 | - | - | - | - |
| 8. Glucose | 9.8 (0.3) | 5.2 (0.9) | 23 | 2612 | - | - | 10 | 1292 | 13 | 1323 | - | - | - | - |
| 9. Insulin | 6.2 (0.5) | 19.9 (14.5) | 26 | 2850 | - | - | 10 | 1463 | 17 | 1387 | - | - | - | - |
| **9. G21** |  |  |  |  |  |  |  |  |  |  |  |  |  |  |
| 1. SBP | 7.2 (0.2) | 105.5 (8.9) | 92 | 5047 | 1890 | 4727 | 41 | 2615 | 51 | 2432 | 40 | 52 | - | - |
| 2. DBP | 7.2 (0.2) | 70.0 (7.6) | 92 | 5047 | 1890 | 4727 | 41 | 2615 | 51 | 2432 | 40 | 52 | - | - |
| 3. HR | 10.2 (0.3) | 82.7 (10.9) | 92 | 4660 | 1696 | 4369 | 43 | 2390 | 49 | 2270 | 37 | 55 | - | - |
| 4. TC | 7.2 (0.2) | 4.4 (0.7) | 69 | 3972 | 1509 | 3729 | 33 | 2047 | 36 | 1925 | 28 | 41 | - | - |
| 5. HDL-C | 7.2 (0.2) | 1.5 (0.3) | 69 | 3974 | 1511 | 3730 | 33 | 2049 | 36 | 1925 | 28 | 41 | - | - |
| 6. LDL-C | 7.2 (0.2) | 2.6 (0.6) | 69 | 3954 | 1498 | 3712 | 33 | 2036 | 36 | 1918 | 28 | 41 | - | - |
| 7. TG | 7.2 (0.2) | 0.8 (0.4) | 69 | 3974 | 1511 | 3730 | 33 | 2049 | 36 | 1925 | 28 | 41 | - | - |
| 8. Glucose | 7.2 (0.2) | 4.6 (0.4) | 69 | 3973 | 1510 | 3729 | 33 | 2049 | 36 | 1924 | 28 | 41 | - | - |
| 9. Insulin | 7.2 (0.2) | 5.6 (4.3) | 65 | 3488 | 1330 | 3278 | 32 | 1788 | 33 | 1700 | 25 | 40 | - | - |
| 10. HbA1c | 7.2 (0.2) | 5.2 (0.4) | 54 | 3143 | 1165 | 2951 | 23 | 1618 | 31 | 1525 | 22 | 32 | - | - |
| **10. GUSTO** |  |  |  |  |  |  |  |  |  |  |  |  |  |  |
| 1. SBP | 3.0 (0.1) | 97.0 (8.4) | 48 | 694 | - | - | 27 | 356 | 20 | 338 | 40 | 7 | 33 | 10 |
| 2. DBP | 3.0 (0.1) | 57.6 (5.8) | 48 | 694 | - | - | 27 | 356 | 20 | 338 | 40 | 7 | 33 | 10 |
| 3. HR | 3.0 (0.1) | 107.5 (11.0) | 48 | 692 | - | - | 27 | 356 | 20 | 336 | 40 | 7 | 33 | 10 |
| 4. TC | 6.1 (4.1) | 4.3 (0.9) | 26 | 363 | - | - | 17 | 177 | 9 | 186 | 22 | 4 | 18 | 6 |
| 5. HDL-C | 6.1 (4.1) | 1.4 (0.3) | 26 | 362 | - | - | 17 | 177 | 9 | 185 | 22 | 4 | 18 | 6 |
| 6. LDL-C | 6.1 (4.1) | 2.5 (0.7) | 26 | 361 | - | - | 17 | 176 | 9 | 185 | 22 | 4 | 18 | 6 |
| 7. TG | 6.1 (4.1) | 0.8 (0.4) | 26 | 363 | - | - | 17 | 177 | 9 | 186 | 22 | 4 | 18 | 6 |
| 8. Glucose | 6.1 (4.1) | 4.5 (0.4) | 30 | 464 | - | - | 19 | 239 | 11 | 225 | 26 | 4 | 22 | 6 |
| 9. Insulin | 6.1 (4.1) | 4.5 (3.2) | 26 | 363 | - | - | 17 | 176 | 9 | 187 | 22 | 4 | 18 | 6 |
| **11. HGS** |  |  |  |  |  |  |  |  |  |  |  |  |  |  |
| 1. SBP | 11.2 (0.7) | 120.6 (13.4) | 60 | 2110 | - | - | 35 | 1039 | 25 | 1071 | 22 | 38 | - | - |
| 2. DBP | 11.2 (0.7) | 69.9 (9.8) | 60 | 2109 | - | - | 35 | 1039 | 25 | 1070 | 22 | 38 | - | - |
| 3. HR | 11.2 (0.7) | 87.1 (12.4) | 58 | 1928 | - | - | 33 | 934 | 25 | 994 | 20 | 38 | - | - |
| 4. TC | 11.2 (0.7) | 4.3 (0.8) | 60 | 2104 | - | - | 34 | 1038 | 26 | 1066 | 21 | 39 | - | - |
| 5. HDL-C | 11.2 (0.7) | 1.5 (0.4) | 60 | 2104 | - | - | 34 | 1038 | 26 | 1066 | 21 | 39 | - | - |
| 6. LDL-C | 11.2 (0.7) | 2.5 (0.7) | 60 | 2104 | - | - | 34 | 1038 | 26 | 1066 | 21 | 39 | - | - |
| 7. TG | 11.2 (0.7) | 0.7 (0.3) | 60 | 2104 | - | - | 34 | 1038 | 26 | 1066 | 21 | 39 | - | - |
| 8. Glucose | 11.2 (0.7) | 5.1 (0.5) | 60 | 2101 | - | - | 34 | 1037 | 26 | 1064 | 21 | 39 | - | - |
| 9. Insulin | 11.2 (0.7) | 11.9 (8.0) | 60 | 2098 | - | - | 34 | 1036 | 26 | 1062 | 21 | 39 | - | - |
| **12. HUNT** |  |  |  |  |  |  |  |  |  |  |  |  |  |  |
| 1. SBP | 16.0 (1.7) | 122.9 (11.1) | 43 | 9,616 | - | - | 21 | 4,794 | 22 | 4,822 | 10 | 33 | 41 | 2 |
| 2. DBP | 16.0 (1.7) | 68.5 (8.0) | 43 | 9,616 | - | - | 21 | 4,794 | 22 | 4,822 | 10 | 33 | 41 | 2 |
| **13. Piccolipiù** |  |  |  |  |  |  |  |  |  |  |  |  |  |  |
| 1. SBP | 4.4 (0.3) | 100.9 (11.5) | 43 | 1494 | 140 | 1356 | 17 | 758 | 26 | 736 | - | - | - | - |
| 2. DBP | 4.4 (0.3) | 64.8 (11.3) | 43 | 1494 | 140 | 1356 | 17 | 758 | 26 | 736 | - | - | - | - |
| **14. SWS** |  |  |  |  |  |  |  |  |  |  |  |  |  |  |
| 1. SBP | 9.2 (0.3) | 106.0 (9.6) | 14 | 933 | - | - | 9 | 462 | 5 | 471 | - | - | - | - |
| 2. DBP | 9.2 (0.3) | 56.1 (7.7) | 14 | 933 | - | - | 9 | 462 | 5 | 471 | - | - | - | - |
| 3. HR | 9.2 (0.3) | 73.6 (10.2) | 14 | 875 | - | - | 9 | 434 | 5 | 441 | - | - | - | - |

| **Supplementary Table 2**. Comparison of included participants with those excluded due to missing data in two European cohorts | | |
| --- | --- | --- |
|  | Included | Excluded |
| **ALSPAC cohort** |  |  |
| Maternal age – years [mean (SD)] | 29.7 (4.5) | 27.6 (5.1) |
| Maternal BMI – kg/m^2^ [mean (SD)] | 22.9 (3.7) | 22.9 (4.0) |
| Maternal parity [No. (%)] |  |  |
| 0 | 2672 (45.6) | 3203 (44.1) |
| 1 or more | 3193 (54.4) | 4061 (55.9) |
| Maternal ethnicity [No. (%)] |  |  |
| White | 5763 (98.2) | 6318 (96.6) |
| Non-white | 102 (1.7) | 224 (3.4) |
| Maternal education [No. (%)] |  |  |
| Degree or higher | 981 (16.7) | 629 (9.5) |
| Lower than degree | 4884 (83.3) | 6005 (90.5) |
| Maternal smoking [No. (%)] |  |  |
| no | 253 (4.3) | 478 (8.9) |
| yes | 5612 (95.7) | 5477 (92.0) |
| **ABCD cohort** |  |  |
| Maternal age – years [mean (SD)] | 31.9 (4.5) | 30.1 (5.5) |
| Maternal BMI – kg/m^2^ [mean (SD)] | 22.7 (3.9) | 23.2 (4.2) |
| Maternal parity [No. (%)] |  |  |
| 0 | 1584 (55.4) | 2997 (55.4) |
| 1 or more | 1276 (44.6) | 2409 (44.6) |
| Maternal ethnicity [No. (%)] |  |  |
| White | 2293 (80.2) | 3120 (57.9) |
| Non-white | 567 (19.8) | 2272 (42.1) |
| Maternal education [No. (%)] |  |  |
| 7 or more years | 2310 (80.8) | 3397 (63.8) |
| Less than 7 years | 550 (19.2) | 1926 (36.2) |
| Maternal smoking [No. (%)] |  |  |
| no | 2168 (75.8) | 4071 (75.6) |
| Yes | 692 (24.2) | 1317 (24.4) |

Data shows maternal characteristics for those included in the pooled analysis for systolic blood pressure, and for those that were excluded due to missing data on the mode conception, confounders, or systolic blood pressure.

| **Supplementary Table 3**. Results of sub-group meta-analysis by offspring mean age | | | | |
| --- | --- | --- | --- | --- |
| **C-M outcome** | **N-studies** | **N-total** | **N-ART** | **Estimate (95% CI)** |
| **SBP (SD difference)** |  |  |  |  |
| <10 yrs. old | 11 | 23705 | 364 | 0.0 (-0.2 to 0.2) |
| ≥10 yrs. old | 8 | 26493 | 440 | -0.1 (-0.2 to 0.1) |
| ≥10 yrs. old (excl. HUNT & CHART) | 6 | 16631 | 267 | -0.2 (-0.2 to 0.1) |
| **DBP (SD difference)** |  |  |  |  |
| <10 yrs. old | 11 | 23705 | 364 | 0.0 (-0.1 to 0.1) |
| ≥10 yrs. old | 8 | 26488 | 440 | -0.1 (-0.4 to 0.2) |
| ≥10 yrs. old (excl. HUNT & CHART) | 6 | 16359 | 267 | 0.0 (-0.2 to 0.2) |
| **HR (SD difference)** |  |  |  |  |
| <10 yrs. old | 9 | 16449 | 269 | 0.1 (-0.1 to 0.2) |
| ≥10 yrs. old | 7 | 17612 | 393 | 0.0 (-0.1 to 0.1) |
| ≥10 yrs. old (excl. CHART) | 6 | 17405 | 265 | 0.0 (-0.1 to 0.1) |
| **TC (% difference)** |  |  |  |  |
| <10 yrs. old | 8 | 14956 | 212 | 3.3 (0.4 to 6.1) |
| ≥10 yrs. old | 6 | 11890 | 304 | 0.3 (-2.6 to 3.3) |
| ≥10 yrs. old (excl. CHART) |  | 11691 | 176 | -0.4 (-2.9 to 2.0) |
| **HDLc (% difference)** |  |  |  |  |
| <10 yrs. old | 8 | 14959 | 212 | 3.4 (0.7 to 6.2) |
| ≥10 yrs. old | 6 | 11890 | 304 | 5.2 (1.2 to 9.3) |
| ≥10 yrs. old (excl. CHART) |  | 11691 | 176 | 5.6 (0.9 to 10.3) |
| **LDLc (% difference)** |  |  |  |  |
| <10 yrs. old | 7 | 15178 | 205 | 4.2 (-1.1 to 9.6) |
| ≥10 yrs. old | 6 | 12279 | 304 | 0.4 (-5.1 to 6.0) |
| ≥10 yrs. old (excl. CHART) | 5 | 12080 | 176 | -1.0 (-6.9 to 4.9) |
| **TG (% difference)** |  |  |  |  |
| <10 yrs. old | 6 | 14591 | 198 | 0.3 (0.3 to -5.0) |
| ≥10 yrs. old | 6 | 12291 | 304 | -7.4 (-14.1 to -0.8) |
| ≥10 yrs. old (excl. CHART) |  | 12092 | 176 | -9.1 (-16.1 to -21.7) |
| **Glucose (% difference)** |  |  |  |  |
| <10 yrs. old | 6 | 11736 | 181 | 0.6 (-1.8 to 3.1) |
| ≥10 yrs. old | 6 | 11878 | 299 | -0.2 (-1.8 to 1.3) |
| ≥10 yrs. old (excl. CHART) | 5 | 11685 | 175 | -0.7 (-2.2 to 0.9) |
| **Insulin (% difference)** |  |  |  |  |
| <10 yrs. old | 5 | 11451 | 162 | -3.2 (-14.5 to 8.2) |
| ≥10 yrs. old | 5 | 11056 | 287 | -12.7 (-22.0 to -3.4) |
| ≥10 yrs. old (excl. CHART) | 4 | 10860 | 161 | -16.0 (-25.0 to -7.1) |
| Results show confounder-adjusted pooled mean differences in cardio-metabolic outcomes between ART-conceived and NC offspring, from meta-analysis in sub-groups by offspring’s mean age at outcome assessment. Results shown before and after excluding the two cohorts with incomplete confounders (HUNT and CHART). SBP: systolic blood pressure, DBP: diastolic blood pressure, HR: heart rate, TC: total cholesterol. HDLc: high-density lipoprotein, LDLc low-density lipoprotein cholesterol, TG: triglycerides. | | | | |

| **Supplementary Table 4**. Predicted mean difference in cardio-metabolic outcomes from childhood to adulthood between ART-conceived and NC offspring, from trajectory models in four birth cohort studies | |
| --- | --- |
|  | **Predicted mean difference for ART minus NC (95%CI)** |
| **SBP (difference in mmHg)** |  |
| age 4 years | -1.23 (-2.25 to -0.2) |
| age 6 years | -1.26 (-2.19 to -0.34) |
| age 8 years | -0.8 (-1.66 to 0.07) |
| age 10 years | -0.13 (-1.2 to 0.93) |
| age 12 years | -0.05 (-1.15 to 1.05) |
| age 14 years | -0.05 (-1.49 to 1.39) |
| age 16 years | 0.27 (-1.47 to 2.02) |
| age 18 years | 0.88 (-0.96 to 2.72) |
| age 20 years | 1.72 (-0.16 to 3.59) |
| age 22 years | 2.73 (0.67 to 4.78) |
| age 24 years | 3.86 (1.32 to 6.4) |
| age 26 years | 5.06 (1.76 to 8.35) |
| **DBP (difference in mmHg)** |  |
| age 4 years | -0.27 (-1.08 to 0.54) |
| age 6 years | -1.26 (-1.99 to -0.54) |
| age 8 years | -1.19 (-1.86 to -0.51) |
| age 10 years | -0.36 (-1.21 to 0.49) |
| age 12 years | -0.05 (-0.92 to 0.82) |
| age 14 years | -0.01 (-1.17 to 1.15) |
| age 16 years | 0.09 (-1.32 to 1.51) |
| age 18 years | 0.27 (-1.23 to 1.77) |
| age 20 years | 0.5 (-1.02 to 2.02) |
| age 22 years | 0.77 (-0.91 to 2.46) |
| age 24 years | 1.08 (-1.03 to 3.19) |
| age 26 years | 1.4 (-1.36 to 4.15) |
| **HR (difference in bpm)** |  |
| age 4 years | 1.61 (0.22 to 3.01) |
| age 6 years | 0.69 (-0.45 to 1.84) |
| age 8 years | 0.12 (-0.93 to 1.16) |
| age 10 years | 0.1 (-1.15 to 1.35) |
| age 12 years | 0.74 (-0.58 to 2.06) |
| age 14 years | 1.45 (-0.26 to 3.16) |
| age 16 years | 1.84 (-0.24 to 3.93) |
| age 18 years | 1.95 (-0.26 to 4.15) |
| age 20 years | 1.82 (-0.4 to 4.04) |
| age 22 years | 1.51 (-0.88 to 3.9) |
| age 24 years | 1.08 (-1.84 to 4) |
| age 26 years | 0.58 (-3.19 to 4.35) |
| **TC (difference in mmol/l)** |  |
| age 4 years | -0.03 (-0.19 to 0.13) |
| age 6 years | 0.06 (-0.03 to 0.15) |
| age 8 years | 0.09 (0 to 0.18) |
| age 10 years | 0.04 (-0.04 to 0.12) |
| age 12 years | -0.01 (-0.1 to 0.09) |
| age 14 years | -0.03 (-0.14 to 0.08) |
| age 16 years | -0.05 (-0.16 to 0.07) |
| age 18 years | -0.05 (-0.17 to 0.07) |
| age 20 years | -0.05 (-0.17 to 0.09) |
| age 22 years | -0.03 (-0.18 to 0.12) |
| age 24 years | -0.01 (-0.2 to 0.18) |
| age 26 years | 0.01 (-0.23 to 0.27) |
| **HDLc (difference in mmol/l)** |  |
| age 4 years | -0.03 (-0.12 to 0.07) |
| age 6 years | 0.02 (-0.03 to 0.07) |
| age 8 years | 0.06 (0.01 to 0.1) |
| age 10 years | 0.08 (0.04 to 0.12) |
| age 12 years | 0.08 (0.03 to 0.14) |
| age 14 years | 0.08 (0.01 to 0.14) |
| age 16 years | 0.06 (0 to 0.13) |
| age 18 years | 0.04 (-0.03 to 0.11) |
| age 20 years | 0.01 (-0.06 to 0.08) |
| age 22 years | -0.03 (-0.1 to 0.06) |
| age 24 years | -0.07 (-0.16 to 0.03) |
| age 26 years | -0.11 (-0.23 to 0.01) |
| **LDLc (difference in mmol/l)** |  |
| age 4 years | -0.01 (-0.11 to 0.11) |
| age 6 years | 0.02 (-0.05 to 0.09) |
| age 8 years | 0.01 (-0.07 to 0.09) |
| age 10 years | -0.04 (-0.11 to 0.03) |
| age 12 years | -0.08 (-0.16 to 0.01) |
| age 14 years | -0.09 (-0.19 to 0.01) |
| age 16 years | -0.1 (-0.2 to 0.01) |
| age 18 years | -0.09 (-0.19 to 0.02) |
| age 20 years | -0.07 (-0.18 to 0.05) |
| age 22 years | -0.05 (-0.18 to 0.09) |
| age 24 years | -0.01 (-0.18 to 0.17) |
| age 26 years | 0.03 (-0.19 to 0.29) |
| **TG (difference in mmol/l)** |  |
| age 4 years | 0.07 (-0.04 to 0.2) |
| age 6 years | 0.03 (-0.02 to 0.09) |
| age 8 years | 0 (-0.05 to 0.05) |
| age 10 years | -0.03 (-0.07 to 0.02) |
| age 12 years | -0.04 (-0.09 to 0.02) |
| age 14 years | -0.04 (-0.1 to 0.03) |
| age 16 years | -0.03 (-0.1 to 0.05) |
| age 18 years | -0.01 (-0.08 to 0.07) |
| age 20 years | 0.01 (-0.06 to 0.09) |
| age 22 years | 0.04 (-0.04 to 0.14) |
| age 24 years | 0.08 (-0.03 to 0.21) |
| age 26 years | 0.12 (-0.02 to 0.29) |
| **glucose (difference in mmol/l)** |  |
| age 4 years | 0.09 (0 to 0.19) |
| age 6 years | 0.04 (-0.01 to 0.1) |
| age 8 years | -0.01 (-0.05 to 0.03) |
| age 10 years | -0.04 (-0.1 to 0.01) |
| age 12 years | -0.07 (-0.14 to 0) |
| age 14 years | -0.08 (-0.16 to 0) |
| age 16 years | -0.08 (-0.16 to 0) |
| age 18 years | -0.07 (-0.16 to 0.02) |
| age 20 years | -0.05 (-0.15 to 0.05) |
| age 22 years | -0.02 (-0.15 to 0.11) |
| age 24 years | 0.01 (-0.15 to 0.18) |
| age 26 years | 0.05 (-0.15 to 0.26) |
| Data shows predicted mean differences (and 95%CIs) in cardio-metabolic outcomes from childhood to adulthood between ART-conceived and NC offspring. Results obtained from multicohort natural cubic spline mixed effects trajectory models in ALSPAC, G21, ABCD, and GUSTO cohorts, that were adjusted for offspring sex, maternal age, parity, BMI, smoking, education, and ethnicity. All models included an interaction between ART and age. SBP: systolic blood pressure, DBP: diastolic blood pressure, HR: heart rate, TC: total cholesterol, HDLc: high-density lipoprotein cholesterol, LDLc: low-density lipoprotein cholesterol, TG: triglycerides | |

### **Supplementary Figure 1**. Directed Acyclic Graph used to identify potential confounders

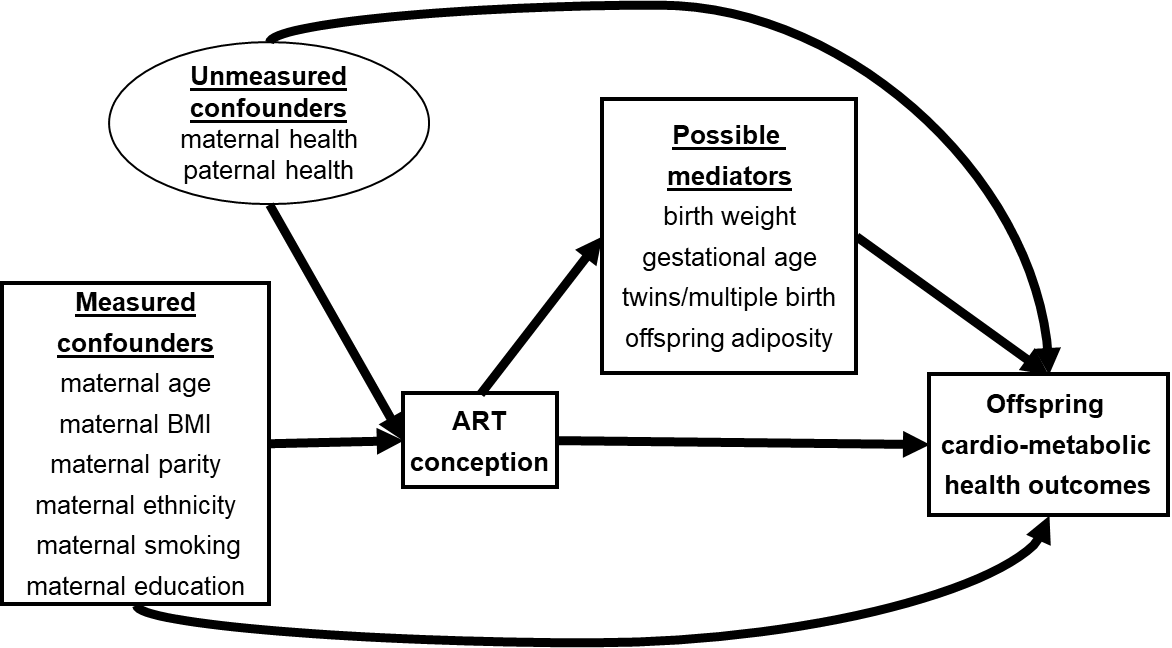

### **Supplementary Figure 2**. Cohort-specific results for the mean differences in cardio-metabolic outcomes between ART and NC offspring

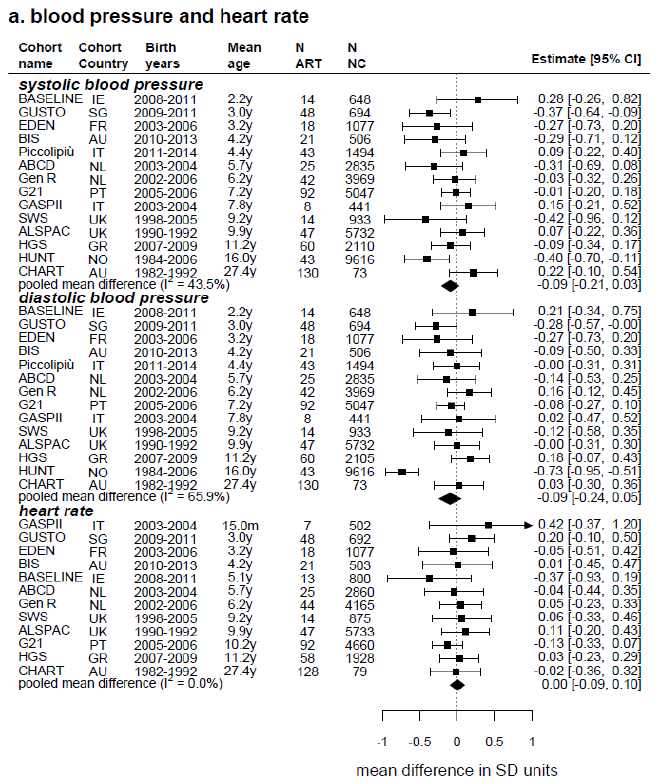

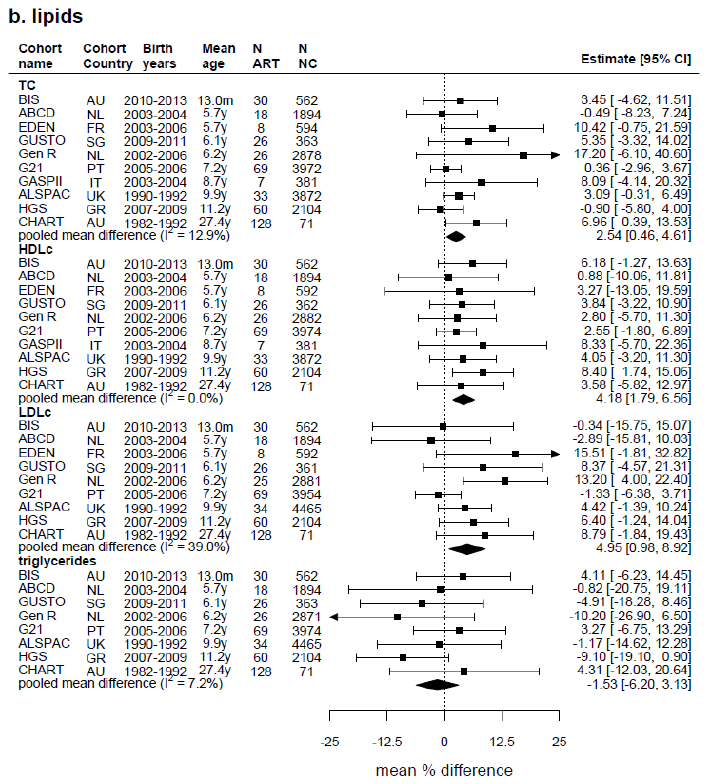

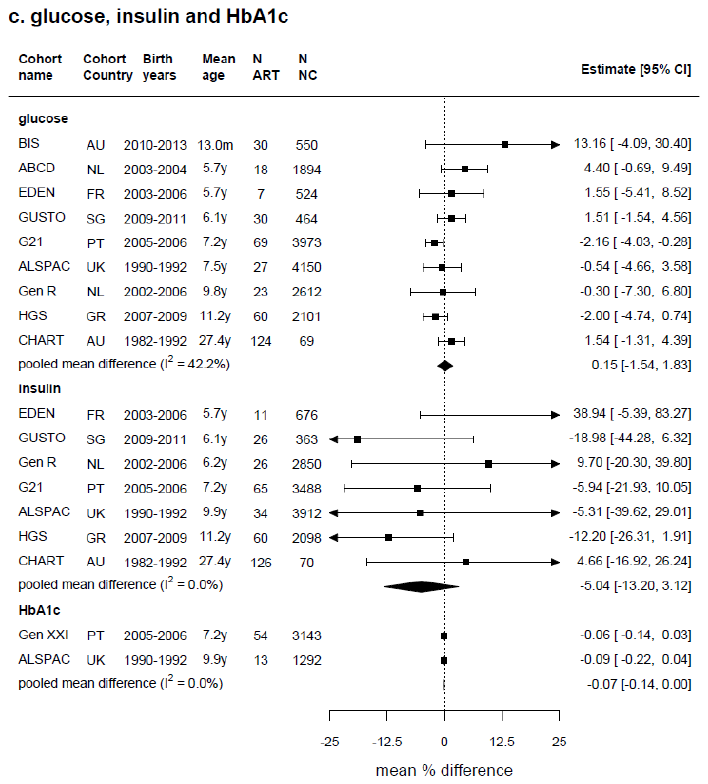

Estimates show confounder-adjusted mean differences in SD units [and 95% confidence intervals] in each cardio-metabolic outcome between ART-conceived and NC offspring in each cohort separately and combined (using random-effects meta-analysis). Cohort specific models were adjusted as fully as possible for maternal age, parity, BMI, smoking, education, and ethnicity, plus offspring age and sex. SBP: systolic blood pressure, DBP: diastolic blood pressure, HR: heart rate, TC: total cholesterol, HDLc: high-density lipoprotein cholesterol, LDLc: low-density lipoprotein cholesterol, TG: triglycerides, HbA1c: glycated haemoglobin.

### **Supplementary Figure 3**. Leave 1 out analysis results

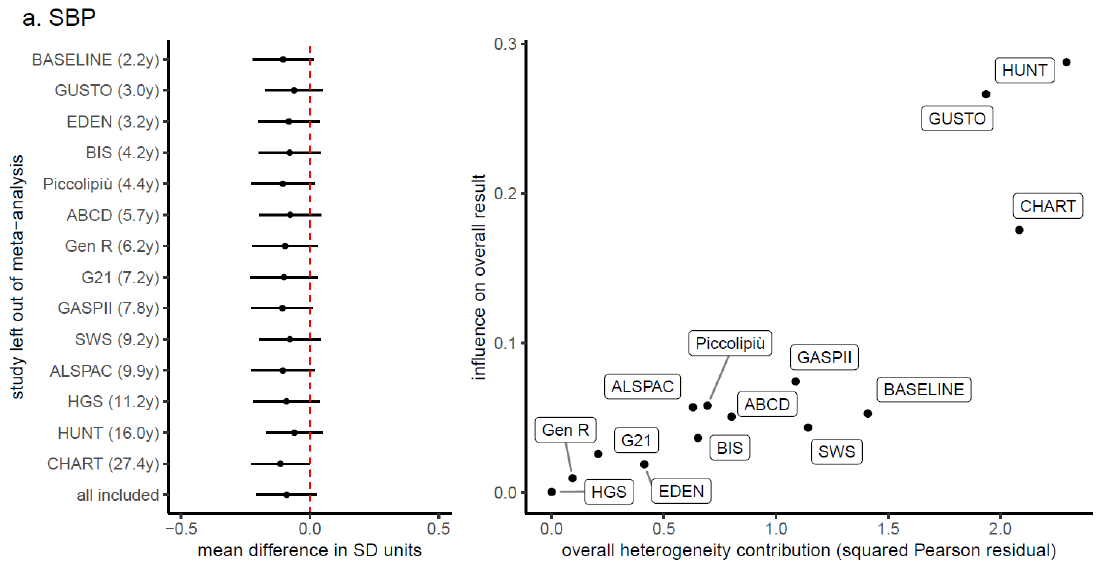

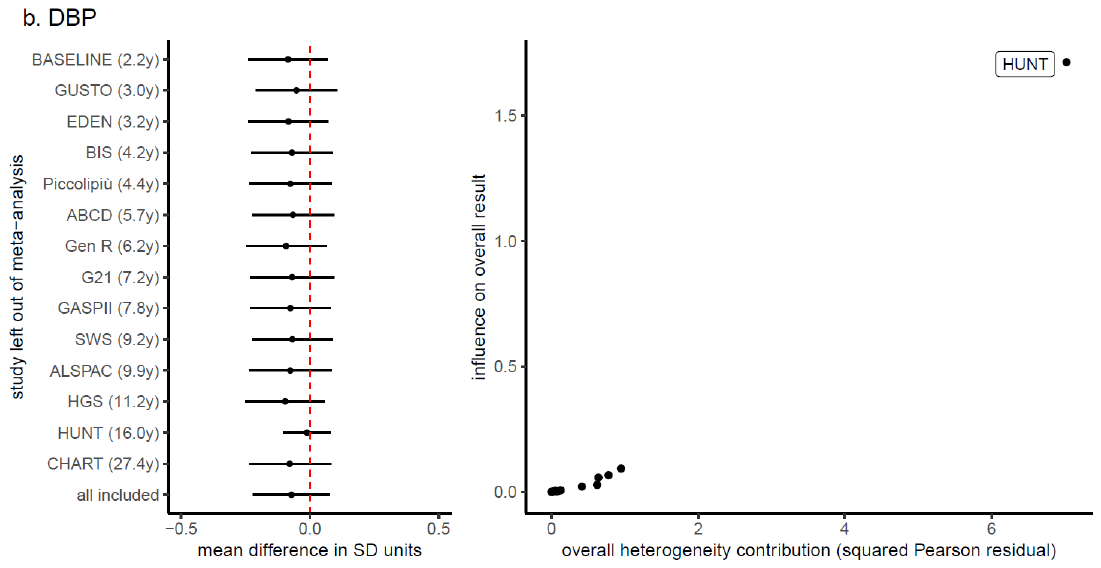

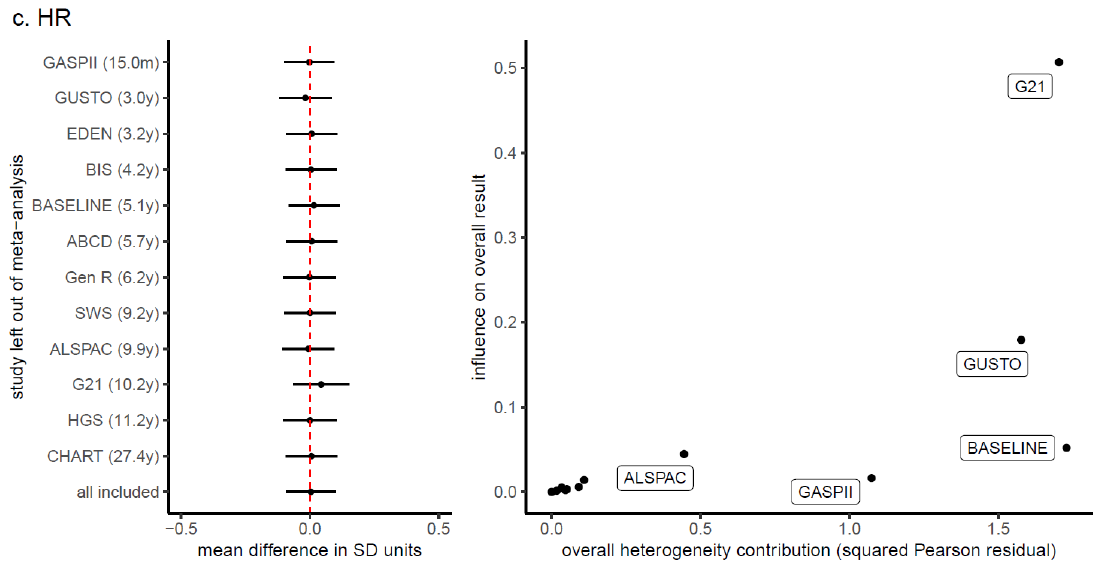

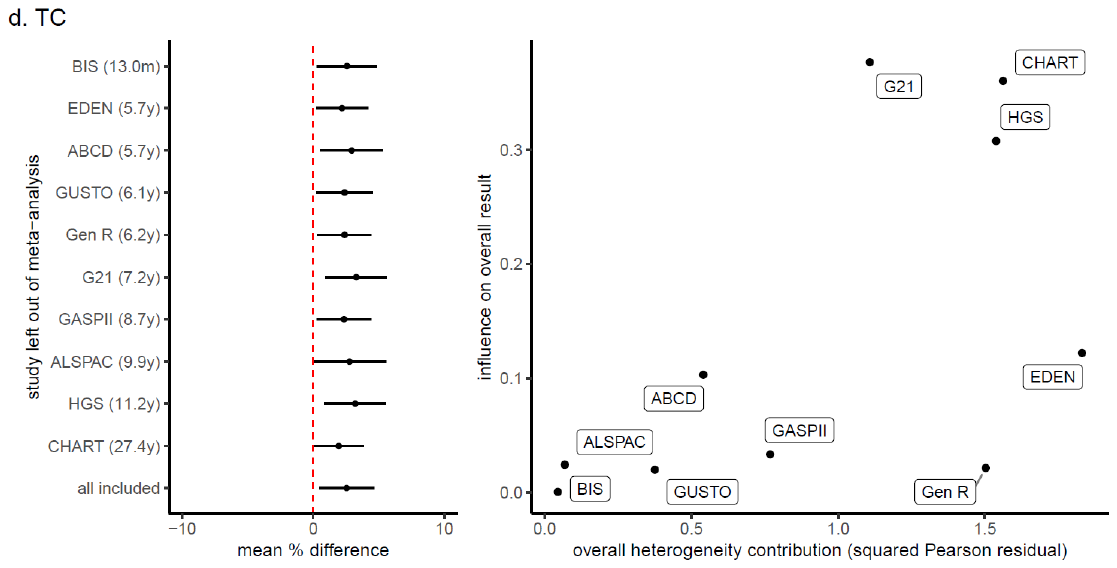

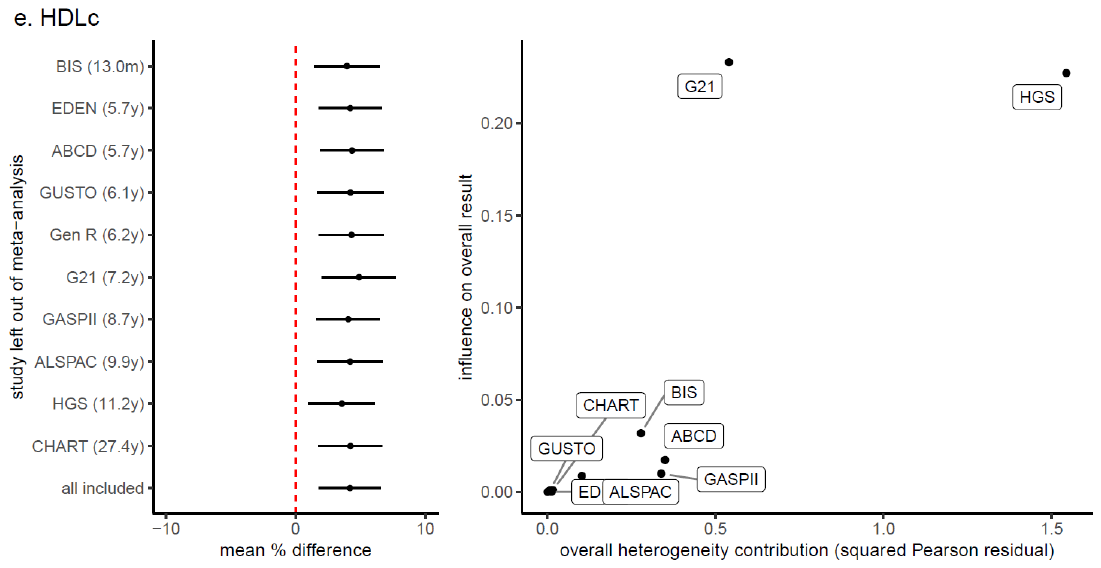

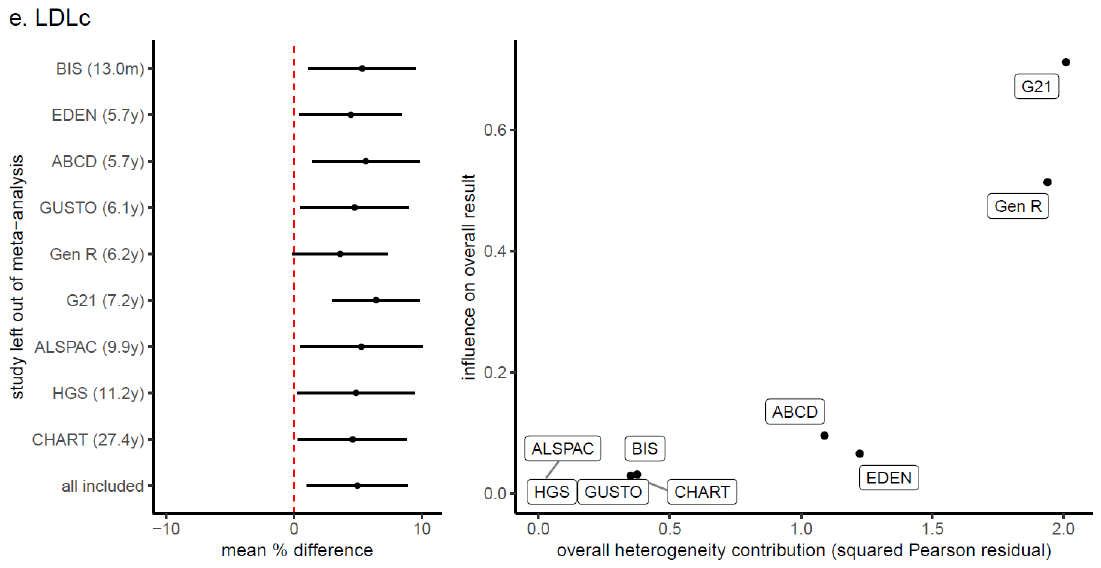

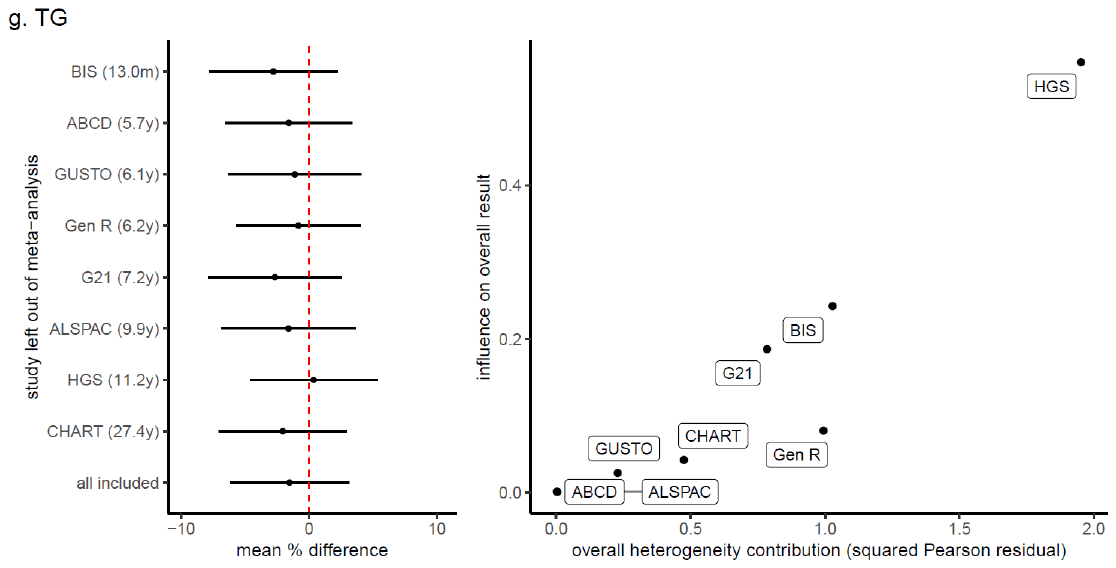

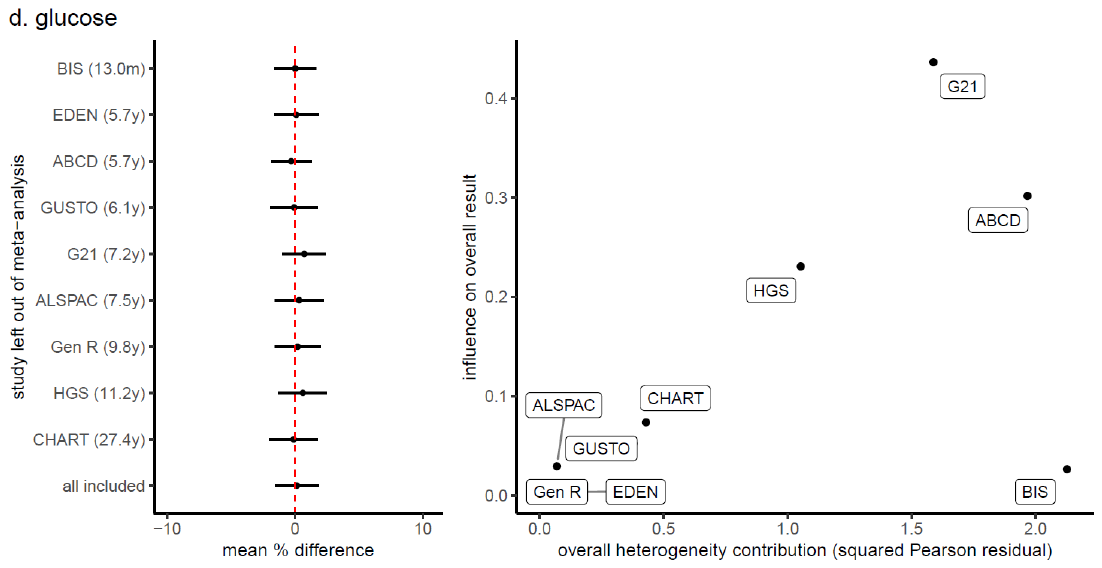

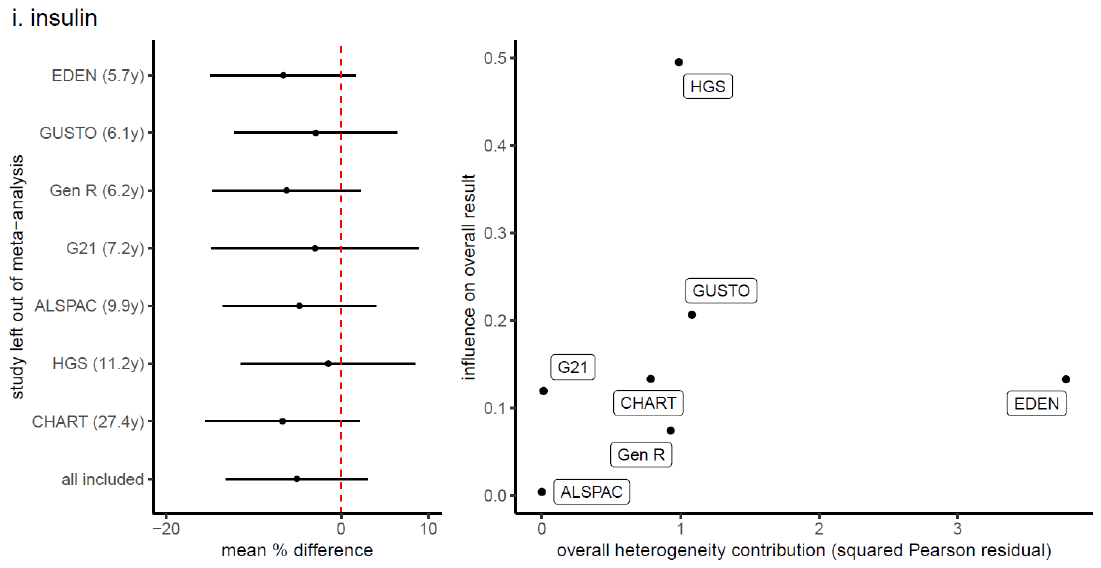

The left panels show the pooled estimates when leaving out each study in turn, as well the estimate for all studies combined (i.e., the main results from Figure 1). Right panels show a modified Baujat plot where the x-axis corresponds to the squared Pearson residual of a study, and y-axis reflects the standardized squared difference between the fitted value for the study with and without the study included in the model fitting. This plot shows the contribution of each study to the overall heterogeneity (x-axis) as well as its effect on the summary (pooled) estimate (y-axis).

### **Supplementary Figure 4**. Mean difference in cardio-metabolic outcomes between ART-conceived and NC offspring, comparing results in all participants to singleton births only

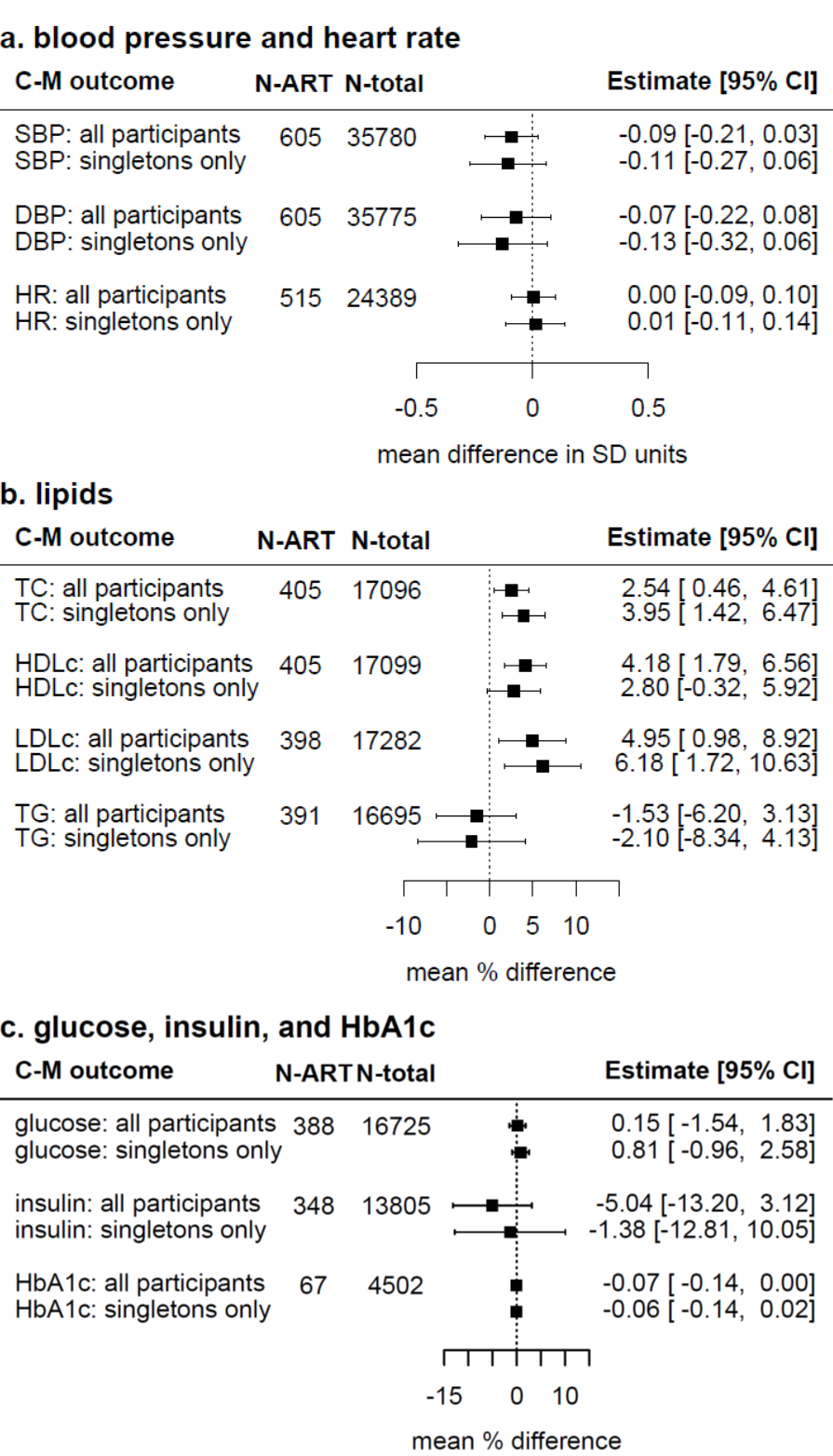

Results show the confounder-adjusted pooled mean differences in SD units and 95% confidence intervals in cardio-metabolic outcomes between ART-conceived and NC offspring (ART minus NC), comparing results in all participants (i.e., those presented in Figures 1) to singleton birth offspring. Cohort-specific estimates were adjusted (as fully as possible) for maternal age, parity, BMI, smoking, education, ethnicity (or country of birth), plus offspring sex and age at outcome assessment. Of the total 14 cohorts in this study, 9 included both singletons and multiple births, and 5 cohorts included singletons births only.

### **Supplementary Figure 5**. Mean difference in cardio-metabolic outcomes between ART-conceived and NC offspring, after further adjustment for birth weight, gestational age, and offspring BMI

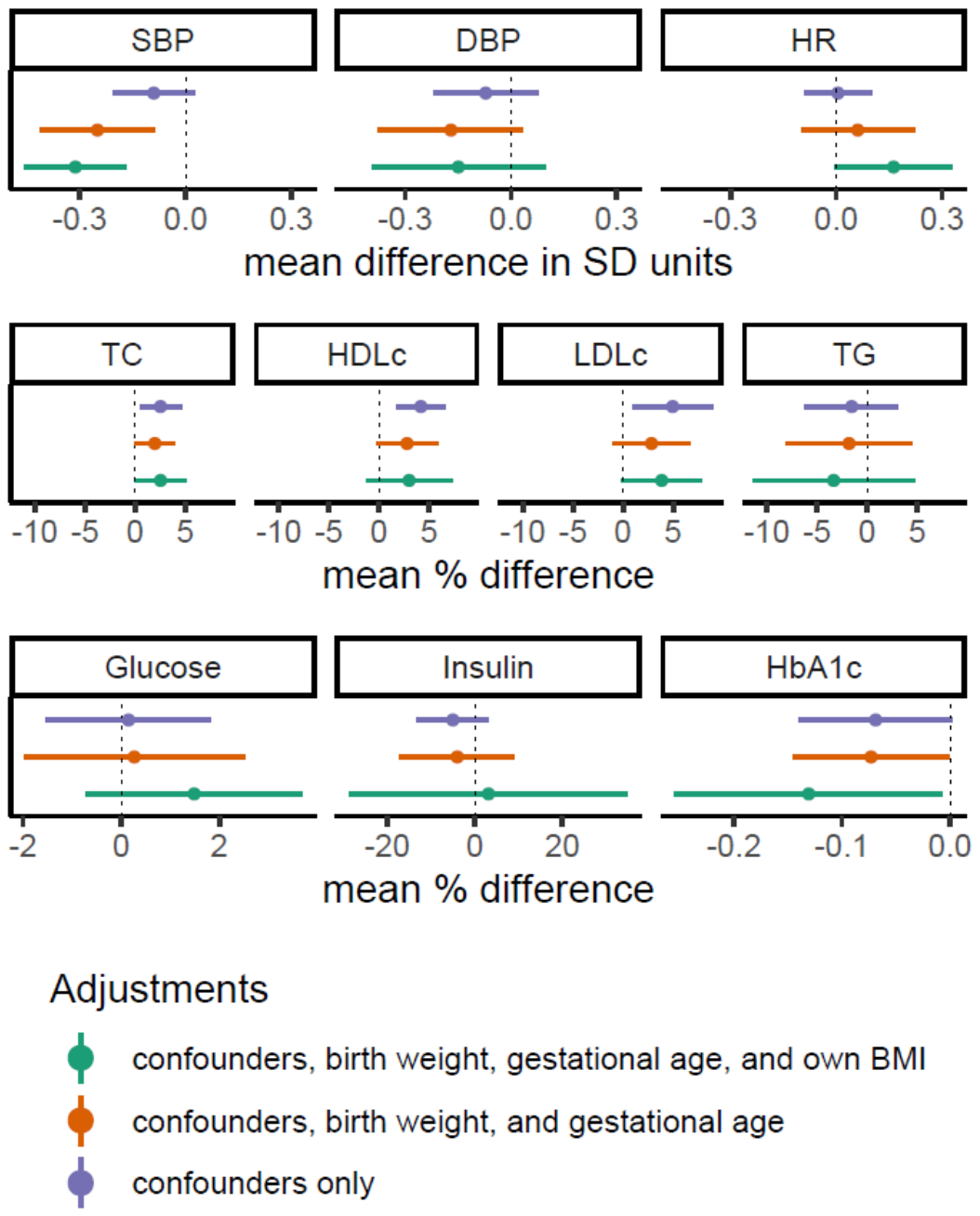

Results show the pooled confounder-adjusted mean differences in SD units and 95% confidence intervals in cardio-metabolic outcomes between ART-conceived and NC offspring (ART minus NC), before and after further adjustment for potential mediators: birthweight, gestational age, and offspring BMI (before or at outcome assessment). Cohort-specific estimates were adjusted (as fully as possible) for maternal age, parity, BMI, smoking, education, ethnicity (or country of birth), plus offspring sex and age at outcome assessment.

### **Supplementary Figure 6**. Predicted mean cardio-metabolic trajectories from age 3-26 years in ART-conceived and NC offspring, in singleton births only

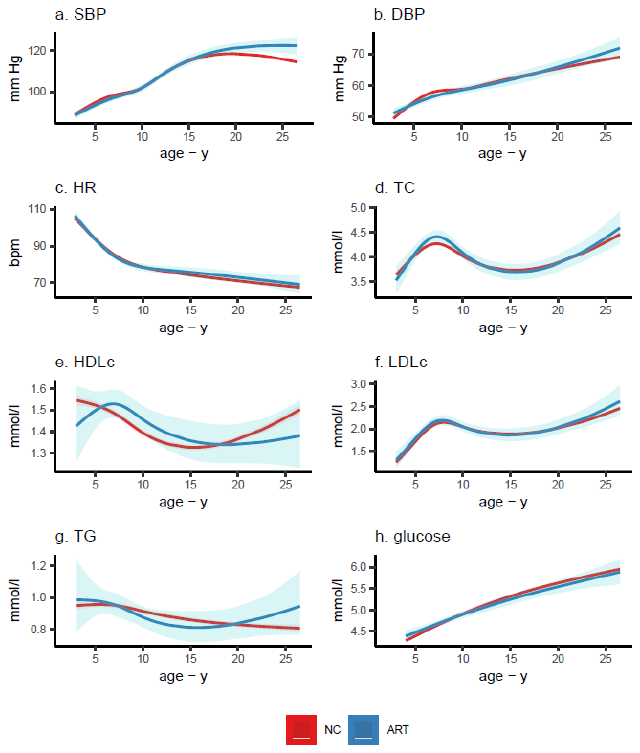

Figure shows the predicted mean trajectories in cardio-metabolic outcomes from childhood to adulthood for ART-conceived and NC singleton offspring. Predicted means were obtained from multicohort (ALSPAC, G21, ABCD, and GUSTO cohorts) natural cubic spline mixed effects trajectory models. Models were adjusted for offspring sex, maternal age, parity, BMI, smoking, education, and ethnicity, and included an interaction between ART and age. SBP: systolic blood pressure, DBP: diastolic blood pressure, HR: heart rate, TC: total cholesterol, HDLc: high-density lipoprotein cholesterol, LDLc: low-density lipoprotein cholesterol, TG: triglycerides. The numbers of offspring included in this analysis were 14,115 (184 ART) for SBP and DBP, 13,693 (183 ART) for HR, 11,145 (140 ART) for lipids, and 10,937 (139 ART) for glucose.

### **Supplementary Figure 7**. Predicted mean cardio-metabolic trajectories from age 3-26 years in ART-conceived and NC offspring, after further adjustment for birth weight and gestational age

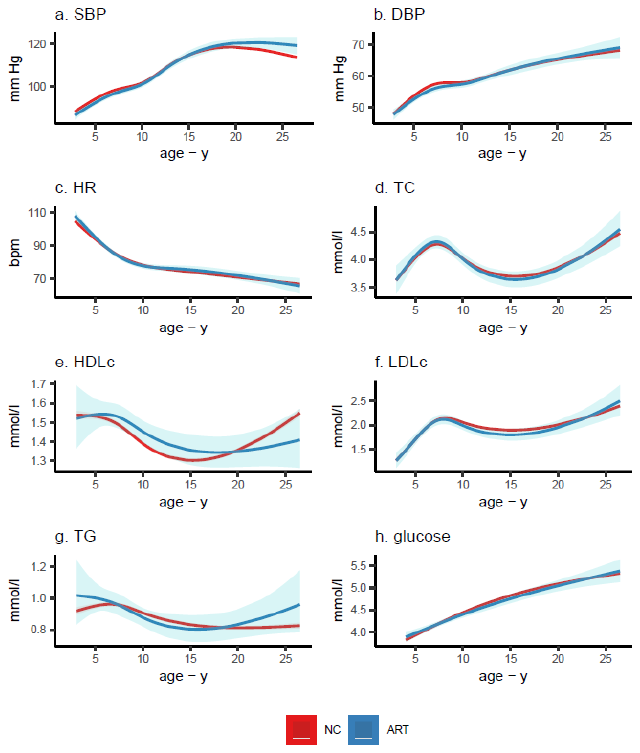

Figure shows the predicted mean trajectories in cardio-metabolic outcomes from childhood to adulthood for ART-conceived and NC offspring, after further adjustment for birth weight and gestational age. Predicted means were obtained from multicohort (ALSPAC, G21, ABCD, and GUSTO cohorts) natural cubic spline mixed effects trajectory models. Models were adjusted for offspring sex, maternal age, parity, BMI, smoking, education, ethnicity, birth weight and gestational age, and included an interaction between ART and age. SBP: systolic blood pressure, DBP: diastolic blood pressure, HR: heart rate, TC: total cholesterol, HDLc: high-density lipoprotein cholesterol, LDLc: low-density lipoprotein cholesterol, TG: triglycerides. Numbers of offspring included in this analysis were 17,114 (240 ART) for SBP and DBP, 16,690 (239 ART) for HR, 13,599 (176 ART) for lipids, and 13,131 (173 ART) for glucose.

### **Supplementary Figure 8**. Predicted mean cardio-metabolic trajectories from age 3-26 years in ART-conceived and NC offspring, after further adjustment for offspring birth weight, gestational age, and BMI

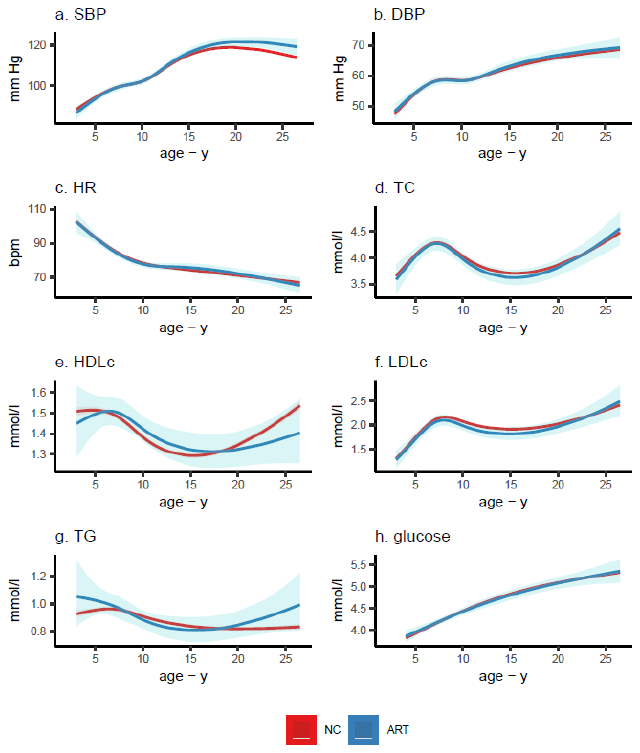

Figure shows the predicted mean trajectories in cardio-metabolic outcomes from childhood to adulthood for ART-conceived and NC offspring, after further adjustment for birth weight, gestational age and offspring BMI Z score. Predicted means were obtained from multicohort (ALSPAC, G21, ABCD, and GUSTO cohorts) natural cubic spline mixed effects trajectory models. Models were adjusted for offspring sex, maternal age, parity, BMI, smoking, education, and ethnicity, birth weight, gestational age, and offspring BMI, and included an interaction between ART and age. SBP: systolic blood pressure, DBP: diastolic blood pressure, HR: heart rate, TC: total cholesterol, HDLc: high-density lipoprotein cholesterol, LDLc: low-density lipoprotein cholesterol, TG: triglycerides. The numbers of offspring included in this analysis were 14,647 (147 ART) for SBP and DBP, 12,155 (147 ART) for HR, 12,002 (123 ART) for lipids, and 11,487 (117 ART) for glucose.
